# Integrating contact tracing and whole-genome sequencing to track the elimination of dog-mediated rabies

**DOI:** 10.1101/2022.11.24.22282675

**Authors:** Kennedy Lushasi, Kirstyn Brunker, Malavika Rajeev, Elaine A Ferguson, Gurdeep Jaswant, Laurie Baker, Roman Biek, Joel Changalucha, Sarah Cleaveland, Anna Czupryna, Anthony R Fooks, Nicodemus Govella, Daniel T Haydon, Paul Johnson, Rudovick Kazwala, Tiziana Lembo, Denise Marston, Msanif Masoud, Mathew Maziku, Eberhard Mbunda, Geofrey Mchau, Ally Z Mohamed, Emmanuel Mpolya, Chanasa Ngeleja, Kija Ng’abhi, Hesron Nonga, Khasim Omar, Kristyna Rysava, Maganga Sambo, Lwitiko Sikana, Rachel Steenson, Katie Hampson

**Author notes:** Deceased (Eberhard Mbunda). Equal contributions (Kennedy Lushasi, Kirstyn Brunker).

## Abstract

**Background:** Dog-mediated rabies is endemic across Africa causing thousands of human deaths annually. A One Health approach to rabies is advocated, comprising emergency post-exposure vaccination of bite victims and mass dog vaccination to break the transmission cycle. However, the impacts and cost-effectiveness of these components are difficult to disentangle.

**Methods:** We combined contact tracing with whole-genome sequencing to track rabies transmission in the animal reservoir and spillover risk to humans from 2010-2020, investigating how the components of a One Health approach reduced the disease burden and eliminated rabies from Pemba island, Tanzania. With the resulting high-resolution spatiotemporal and genomic data we inferred transmission chains, estimated case detection and quantified the public health burden to evaluate these interventions.

**Results:** We resolved five transmission chains co-circulating on Pemba from 2010 that were all eliminated by May 2014. During this period, rabid dogs, human rabies exposures and deaths all progressively declined following initiation and improved implementation of annual islandwide dog vaccination. We identified two introductions to Pemba in late 2016 that seeded re-emergence after dog vaccination had lapsed. The ensuing outbreak was eliminated in October 2018 through reinstated islandwide dog vaccination. While post-exposure vaccines were highly cost-effective ($405 per death averted), their accessibility was limited and only dog vaccination interrupted transmission. A combined One Health approach rapidly eliminated rabies, was highly cost-effective ($1865 per death averted) and saved 20-120 families from rabid dog bites annually.

**Conclusions:** A One Health approach underpinned by dog vaccination is an efficient, cost-effective, equitable and feasible approach to rabies elimination, but needs scaling up across connected populations to sustain the benefits of elimination, as seen on Pemba, and for similar progress to be achieved elsewhere.

**Funding:** Wellcome [207569/Z/17/Z, 095787/Z/11/Z, 103270/Z/13/Z], the UBS Optimus Foundation, and the DELTAS Africa Initiative [Afrique One-ASPIRE/DEL-15-008] comprising a donor consortium of the African Academy of Sciences (AAS), Alliance for Accelerating Excellence in Science in Africa (AESA), the New Partnership for Africa’s Development Planning and Coordinating (NEPAD) Agency, Wellcome [107753/A/15/Z] and the UK government. The rabies elimination demonstration project from 2010-2015 was supported by the Bill & Melinda Gates Foundation (OPP49679) and whole-genome sequencing was partially supported at APHA by Defra grant SE0421.

## Introduction

Every year an estimated 59,000 people die from rabies,^1^ a viral infection transmitted primarily by domestic dogs in low- and middle-income countries (LMICs). While human rabies encephalitis remains incurable, the disease is readily preventable if post-exposure prophylaxis (PEP) is promptly administered to bite victims upon exposure.^2^ Moreover, mass dog vaccination has eliminated dog-mediated rabies from high-income countries and much of the Americas.^3^ Yet, in most African and Asian countries there has been little investment in dog vaccination and rabies circulates unabated^1^, with recommendations to scale up mass dog vaccination.

Although dog vaccination can eliminate dog-mediated rabies, there are challenges to achieving this goal. In most rabies endemic countries in sub-Saharan Africa, dog vaccination campaigns have been sparse and localised.^4^ Moreover, the high reproductive rates and short lifespan of dogs in many LMICs quickly lead to drops in vaccination coverage, with repeat campaigns required to maintain coverage.^5^ The virus can easily spread in dog populations that have low and heterogenous vaccination coverage^6^ and incursions leading to outbreaks are commonly reported,^7–9^ often facilitated by human-mediated movement of dogs incubating infection.^10,11^This situation is compounded by weak surveillance which hinders effective outbreak response and poses a challenge for monitoring progress towards elimination, including how to determine disease freedom.^12^

Across the African continent there are very few documented examples of dog-mediated rabies elimination. We found just five papers reporting four locations on the continent with potential interruption of transmission by dog vaccination; the cities of N’Djamena, Chad^8^ and Harare, Zimbabwe;^13^ Serengeti district in northwest Tanzania,^14^ and KwaZulu-Natal province, South Africa.^15^ In all four locations, endemic circulation has since re-established, with resurgences explained by movement of infected dogs from surrounding areas after dog vaccination campaigns lapsed. The importance of reintroductions in maintaining rabies circulation is further highlighted from long-term surveillance from Bangui, the capital of the Central African Republic^7^ and Serengeti district, Tanzania.^6^ Genomic surveillance can potentially play a role in differentiating rabies introductions from undetected sustained transmission and thus in confirming or refuting rabies elimination and therefore targeting control efforts. However, sequencing of rabies viruses also remains limited in Africa.

Dog-mediated rabies is endemic in East Africa where thousands of human rabies deaths occur each year.^16^ Rabies has circulated on Pemba Island, off mainland Tanzania, since the late 1990s. Dog vaccinations on Pemba first began in 2010, with a small-scale campaign conducted by the animal welfare organisation, World Animal Protection. Over the next five years a rabies elimination demonstration project, funded by the Bill & Melinda Gates Foundation, coordinated by the World Health Organisation and led by the Tanzanian government, was implemented across southeast Tanzania, including Pemba.^17^ Here, we show how these efforts led to rabies elimination, while highlighting how introductions pose challenges to achieving and maintaining rabies-freedom even on a small, relatively isolated, island. Our study is the first to confirm rabies elimination from an African setting, including in response to reintroduction, through quantifying case detection.^6^ Using rigorous contact tracing, we identified chains of transmission within the domestic dog reservoir informed by in-country whole-genome sequencing (the first example in Africa) and cross-species transmission from domestic dogs to humans. This enabled us to fully capture the public health burden and associated cost-effectiveness of both post-exposure vaccination and dog vaccination, as well as their combined use, in achieving and maintaining rabies freedom. Our findings illustrate the critical need to holistically link surveillance with public health and veterinary interventions to cost-effectively reduce the burden of zoonotic pathogens. This case study provides timely lessons given the global strategic plan to eliminate dog-mediated human rabies by 2030.

## Methods

### Epidemiological and laboratory Investigations

Records of bite patients presenting to health facilities were used to initiate contact tracing.^6^ Bite victims and, if known, the owners of biting animals were exhaustively traced, recording all bite incident details, including dates and coordinates. Other people or animals that were identified as bitten were further traced. The status of animals was assessed from their reported behaviour and outcome (whether they died, disappeared or survived), and classified according to WHO case definitions.^4^ Briefly, an animal showing any clinical signs of rabies was considered a suspect case; if a suspect case had a reliable history of contact with a suspect rabid animal and/or was killed, died or disappeared within 10 days of observation of illness, the animal was considered a probable case. Animals that remained alive for more than 10 days after biting a person, were considered healthy. Brain tissue samples were collected from animal carcasses for diagnostic testing whenever possible.^18^

Two batches of sequencing were performed to obtain 16 near whole-genome sequences (WGS) of rabies virus (RABV) from dog brain samples collected on Pemba, with the approach changing as protocols and capacity for in-country sequencing developed.^19^ Eight of these sequences have been previously published within a methods paper^19^ and 8 are published for the first time here. The latter are archived 2011/12 samples (3) and samples (5) from early outbreak surveillance (September/October 2016) that were confirmed RABV positive at Pemba Veterinary Laboratory Department and shipped to the Animal & Plant Health Agency (APHA), UK. Total RNA was extracted using Trizol (Invitrogen) and a real-time PCR assay^20^ was performed to confirm the presence of RABV and indicate viral load. Metagenomic sequencing libraries were prepared and sequenced on an Illumina MiSeq as previously described.^21^ Subsequent sequencing of the 8 additional samples (September 2016 to May 2017) was conducted in-country in August 2017 at the Tanzania Veterinary Laboratory Agency (TVLA) following an end-to-end protocol using a multiplex PCR approach^22^ for MinION (Oxford Nanopore Technology, Oxford, UK) sequencing of RABV genomes.^19^ Fourteen previously unpublished WGS (via the metagenomic approach) from mainland Tanzania (2009 to 2017) are also published here and included in analyses. The newly published sequences are detailed in the appendix (supplementary Table S1).

### Control and prevention measures

We compiled data on rabies control and prevention measures implemented on Pemba. This included the numbers and timing of dog vaccination campaigns, and costs of dog vaccination and PEP provisioning (Supplementary Table S2).

Briefly, the first small-scale dog vaccination campaign (705 dogs vaccinated) on Pemba took place in 2010. This was followed by four annual islandwide campaigns by livestock field officers under Pemba’s department of livestock as part of the elimination demonstration project (Appendix: Supplementary methods) with vaccinations provided free-of-charge.^17^ During the 2013 and 2014 campaigns, dogs were marked with temporary collars upon vaccination and post-vaccination transects carried out in each village (*shehia*) to estimate coverage achieved.^18^

As part of the demonstration project PEP was procured for free provisioning at Pemba’s four district hospitals. Training in administering both intradermal and intramuscular post-exposure vaccination was completed in early 2011. Following the end of the demonstration project in 2015, bite patients were required to pay 30,000 TSh ($12.9) per vial when undergoing post-exposure vaccination, with multiple vials required for a complete PEP course.

In late 2016, a rabies outbreak was detected. The initial government response involved conducting central point dog vaccination campaigns in *shehias* reporting cases. This expanded to island-wide campaigns from 2017 onwards, including door-to-door vaccination in some s*hehias* where dog owners could not bring dogs to allocated central points. In 2017, the government of Zanzibar also began to subsidise PEP, making vaccines free-of-charge at Pemba’s main hospital and in hospitals in Zanzibar (1-day’s ferry travel), otherwise, post-exposure vaccines were available to purchase on the mainland.

### Analyses

Monthly dog populations in each *shehia* were estimated from dog population surveys in 2012, 2017 and 2019 and post-vaccination transects from 2013 and 2014. Vaccination coverage and the level of vaccine-induced immunity were projected monthly from the dog population estimates and campaign records on numbers and timing of dogs vaccinated (Appendix: Supplementary methods).

We compiled costs associated with PEP provisioning and implementing mass dog vaccination on Pemba (Table S2) and estimated expenditure on these interventions from health facility and dog vaccination campaign records over the decade. Cost-effectiveness of PEP was assessed from numbers of rabies-exposed individuals and the probability of rabies progression in the absence of PEP^16^ and for dog vaccination in comparison to human deaths in years without pre-emptive dog vaccination.

Sequence data were used to understand the source and timing of introductions to Pemba and to resolve transmission chains. Raw sequence reads were processed and underwent quality control filtering.^19,21^ Pemba sequences were submitted to RABV-GLUE^23^ to determine which global RABV subclade they belonged to and compared to all publicly available RABV sequence data from the same subclade (Cosmopolitan-AF1b). The genome region and number of sequences varied widely in these data, therefore subsets were used to extract the most relevant for comparison (Appendix: Supplementary methods).

A Bayesian discrete phylogeographic analysis was conducted in BEAST v1.10.4 from a subset of 153 Tanzanian RABV genomes, of which 13 were from the 2016/17 Pemba outbreak. Two independent MCMC chains were run for 250 million steps with an uncorrelated log-normal relaxed molecular clock. Sequences were partitioned into concatenated coding sequence and non-coding sequence, each with a GTR+G substitution model. Two locations were specified for phylogeographic analysis, “Mainland” or “Island” for identifying the source of introductions. Sampled trees were subset to 10,000 trees and summarised as a maximum clade credibility tree, which was examined to determine the timing of introductions. Phylogenies were visualised and annotated in R using the ggtree package.^24^

Transmission trees were reconstructed from the probable case data. Traced progenitors were assigned, otherwise links between cases were inferred from rabies dispersal kernel and serial interval distributions with uncertainties in timings incorporated.^6^ Trees were pruned to distinguish transmission chains and generate sets consistent with the phylogeny (Appendix: Supplementary methods). We estimated case detection from the times between linked cases and the serial interval distribution.^6^ The effective reproduction number R_e_, which describes transmission in the presence of control measures, was calculated from the resulting transmission chains and examined over time and in relation to estimated dog vaccination coverage.

### Ethics

The study was approved by the Zanzibar Medical Research and Ethics Committee (ZAMREC/0001/JULY/014), the Medical Research Coordinating Committee of the National Institute for Medical Research of Tanzania (NIMR/HQ/R.8a/vol.IX/2788), the Ministry of Regional Administration and Local Government (AB.81/288/01), and Ifakara Health Institute Institutional Review Board (IHI/IRB/No:22-2014).

## Results

Rabies was endemic on Pemba in 2010 at the start of the study. That year we traced 32 human rabies exposures, 33 rabid dogs and three human rabies deaths diagnosed from clinical signs and history of exposure (6.77 exposures and 0.63 deaths/ 100,000 persons and 10.5 cases/ 1000 dogs). Initial dog vaccination implemented as part of a rabies elimination demonstration project in 2011 achieved only low and heterogeneous coverage (13% in 2011, ranging from 7-20% across districts), but by 2014 campaigns were island-wide and achieved better coverage (mean 50%, range 46-60%, Figure 1). Correspondingly, human rabies exposures and dog rabies cases declined each year to just 2 each in 2014. The effective reproduction number, R_e_, also declined from ∼1.5 in 2010 to <1 in 2014 (Figure 2). No human rabies exposures, deaths or animal cases were detected from May 2014 until July 2016.

**Figure 1.**
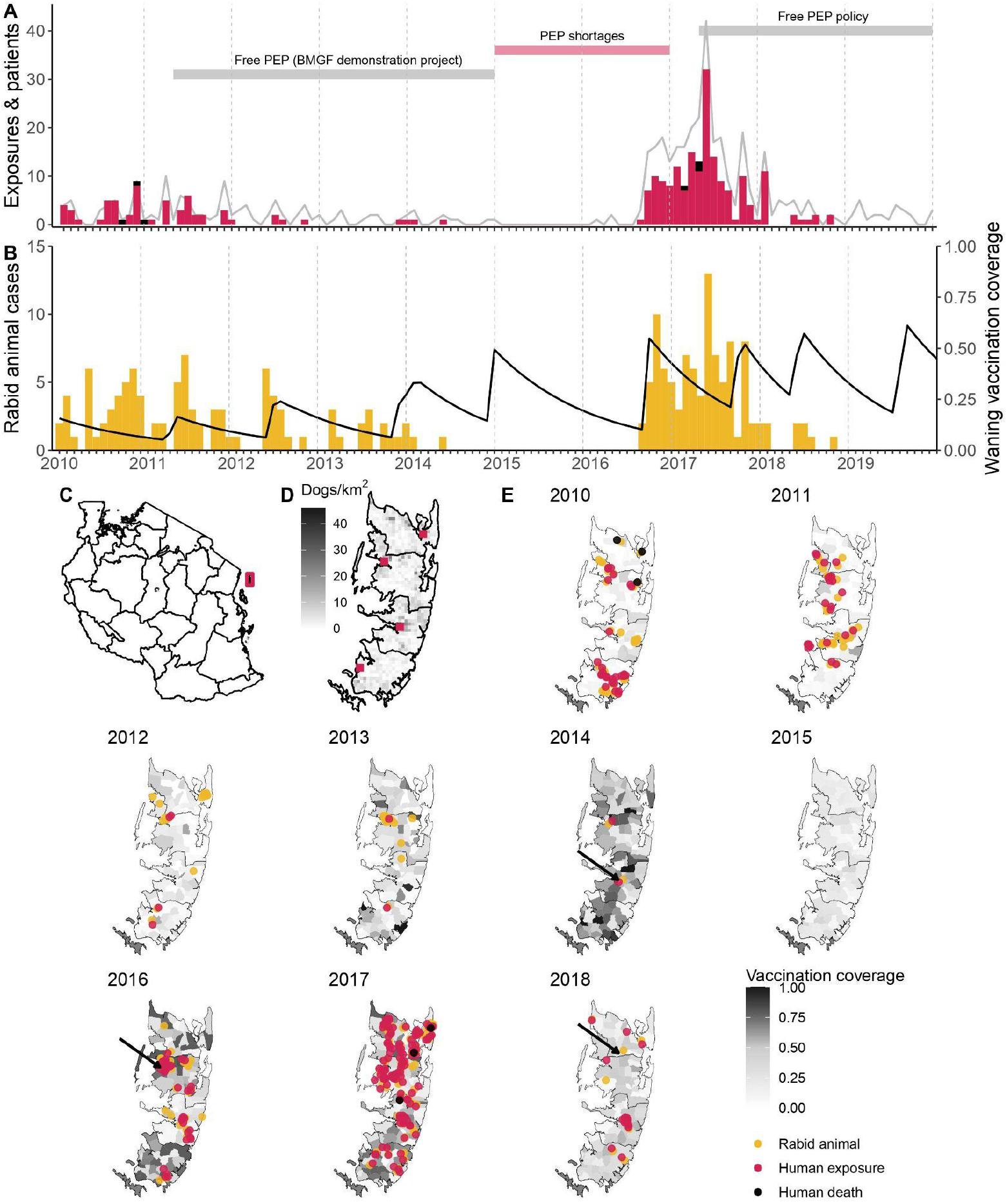
Timeline of rabies on Pemba island in relation to control and prevention measures. **A)** Monthly time series of traced human rabies exposures (red) and deaths (black), and patients presenting to clinics from bites by both healthy and rabid dogs (grey line). Periods when PEP was provided free of charge are indicated by the grey horizontal bars, as well as periods of shortages (red horizontal bar). **B)** Dog rabies cases (orange) in relation to average dog vaccination coverage across the island (black line). **C)** Location of Pemba (red) off the coast of mainland Tanzania. **D)** Density of Pemba’s dog population and location of the four government hospitals that provide PEP (red squares), one in each district. **E)** Dog rabies cases (orange circles) and human rabies exposures (red circles) and deaths (black circles) each year. Shading indicates dog vaccination coverage in December of each year, projected from the timing of *shehia*-level campaigns, dog turnover and a mean vaccine-induced immunity duration of three years. The arrows point to the last detected animal case in 2014, first detection in the 2016 outbreak and the final case found in 2018.

**Figure 2.**
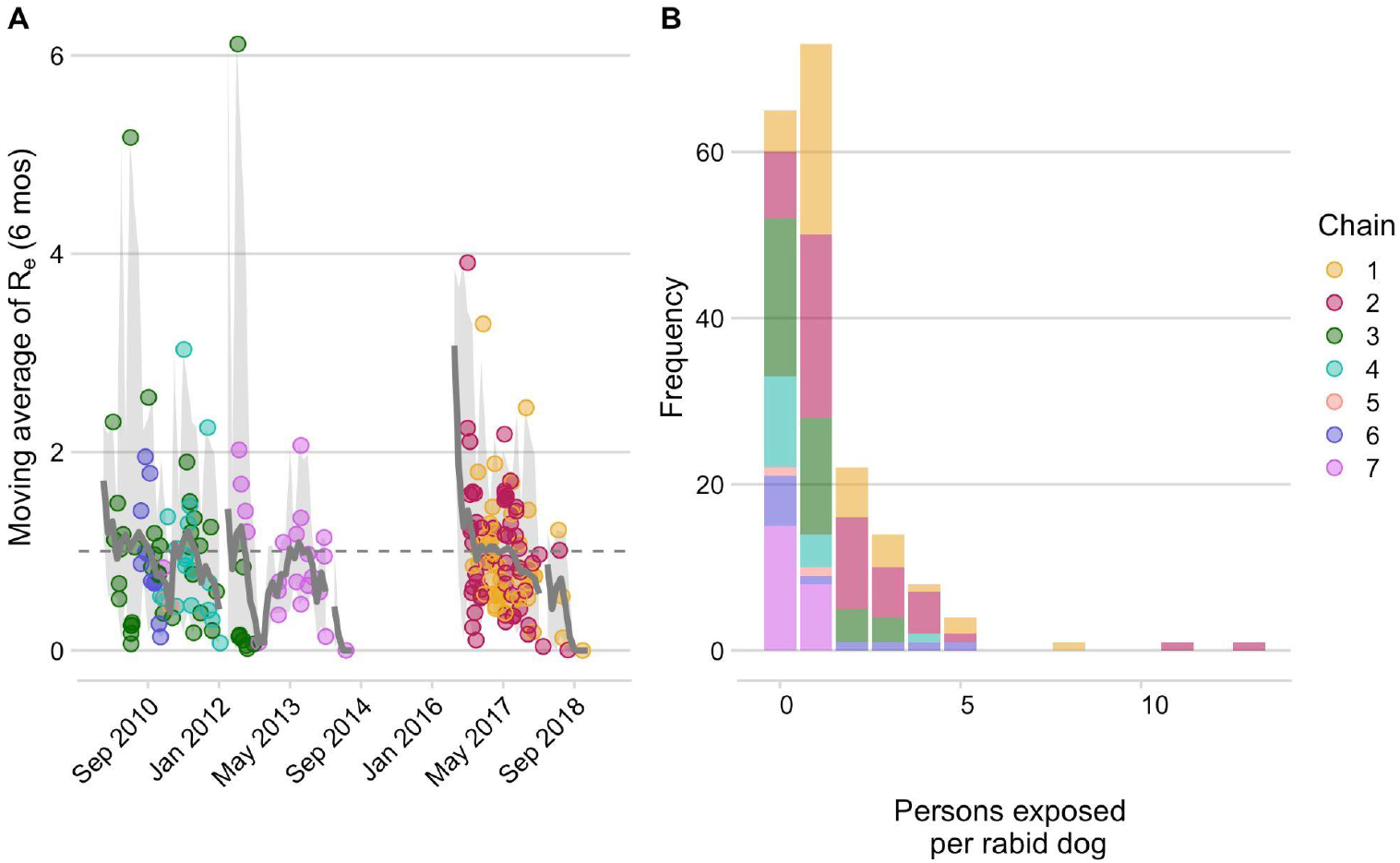
Dog-to-dog rabies transmission and dog-to-human rabies exposures on Pemba. **A)** The effective reproductive number, Re (grey line, moving average over cases in the 2 months prior, the current month, and 3 subsequent months) with 95% quantiles (grey envelope) and mean secondary cases from each traced rabid dog inferred from the bootstrapped transmission trees. **B)** Persons bitten by each traced rabid dog. Points/ bars are coloured by transmission chain (see methods and Figure 4). The grey dashed line indicates an Re equal to 1. In addition to dogs identified as responsible for human exposures shown in B, we estimate around 122 undetected rabid dogs did not bite people or their bite victims did not attend/ were not recorded at health facilities.

In August 2016, an influx of bite patients was seen in hospitals on Pemba. By the year end, we had traced 35 human rabies exposures and 27 dog rabies cases. In response to this outbreak, the Ministry of Livestock and Fisheries Development initiated dog vaccination in *shehias* with recorded dog cases, but because the outbreak spread rapidly, islandwide dog vaccination was reinstated. In 2017, we traced 3 human rabies deaths, 126 rabies exposures and 62 rabid dogs (26.6 exposures and 0.63 deaths/100,000 people, and 19.6 cases/1000 dogs). High vaccination coverage was achieved consecutively over subsequent annual dog vaccination campaigns from 2017 to 2020 (median 61%, range 46-78% in 2019, Figure 1). Incidence rapidly declined from the 2017 peak with 19 human rabies exposures and 8 dog rabies cases detected in 2019. At the start of the outbreak R_e_ was high (>1.5), but subsequently declined to <1, with all transmission interrupted by October 2018 (Figure 2). No human rabies exposures, deaths or rabid dogs have been identified since (as of September 2022).

Phylogenetic analyses indicated considerable viral diversity from 2010-2014 (Figure 3), during which time we resolved five distinct transmission chains (Figure 4, appendix Movie S1), and detected approximately 54% of dog rabies cases circulating on the island (95% credible intervals (95%CI: 46.4-62.0%, 92 of an estimated 171 rabid dogs from 2010-2014, appendix). Viruses sequenced from the outbreak starting in 2016 belonged to two phylogenetic lineages (Figure 3). The time-scaled phylogeny pointed to two independent introductions taking hold and spreading widely, i.e. not continued transmission of viruses circulating previously. Our estimates of case detection were also higher during this outbreak, at 69% (95%CI: 59.4-81.6%) of dog rabies cases (97 out of an estimated 140). Additional phylogenetic analysis of a partial genome sequence dataset that provides wider spatiotemporal context (Appendix) shows that one of these new lineages shares a common ancestor with sequences from Zanzibar, suggesting a possible link between outbreaks on these islands, or a common source of introduction.

**Figure 3.**
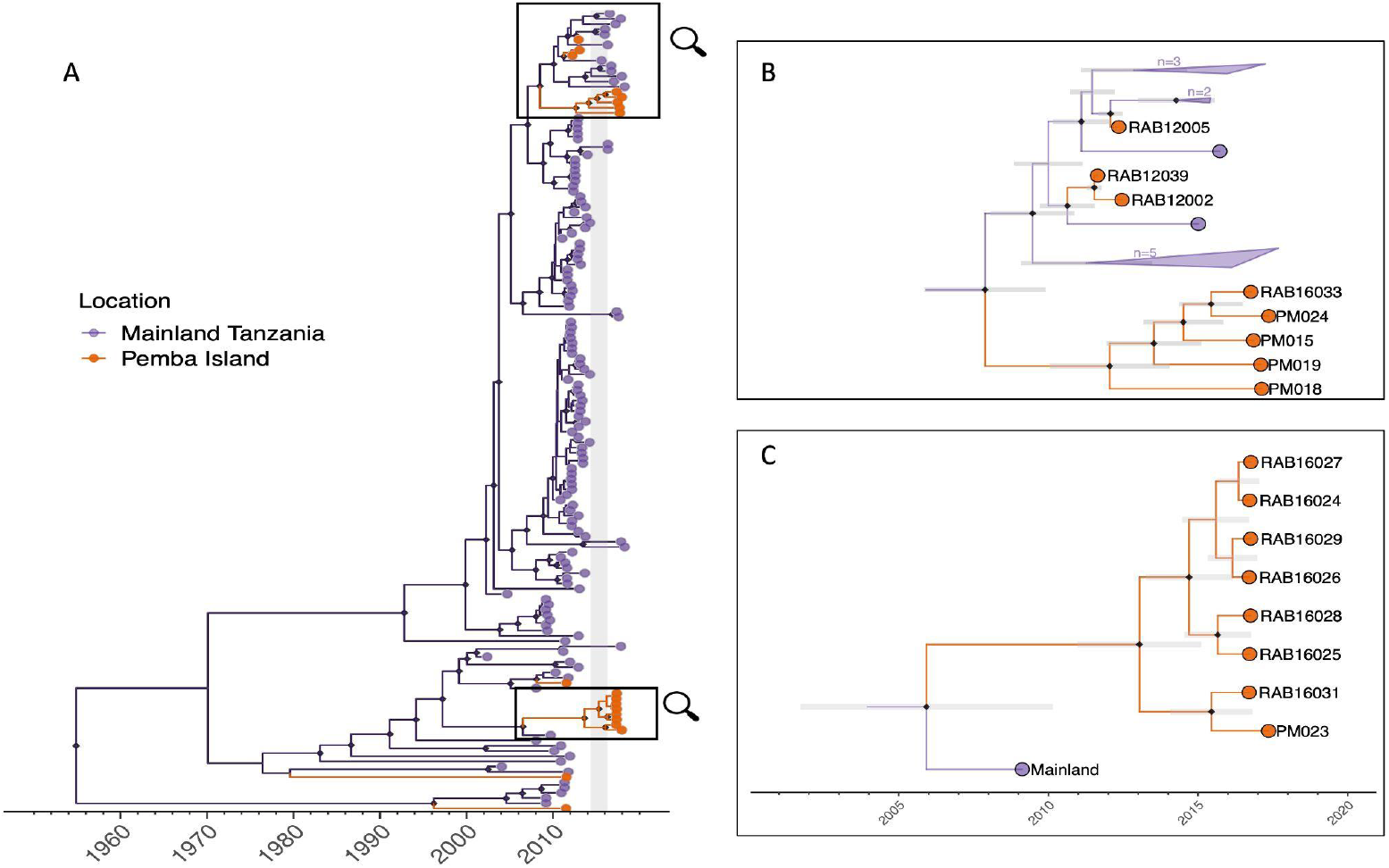
Maximum clade credibility tree (MCC) from discrete phylogeographic analysis to identify rabies virus introductions to Pemba. **A)** Time-calibrated MCC tree of 153 whole-genome sequences from Tanzania, including 13 from the 2016-2018 Pemba outbreak and 6 historical Pemba sequences (2010-2012). Grey vertical bar highlights the window of emergence for the most recent common ancestors of the two introductions that led to the 2016 outbreak (2014.33-2016.29). The expanded subtrees [**B** and **C**] show the Pemba cases one node back from the most recent common ancestor of the 2016 introductions, with branches coloured according to the inferred ancestral location. Black diamonds indicate nodes with >90% posterior support (clade credibilities). Mainland clusters of more than one identical sequence are collapsed. Grey bars represent the 95% highest posterior density interval of node heights, i.e. estimated age of ancestral nodes. Names of sequences are shown so they can be related to metadata Tables (appendix).

**Figure 4.**
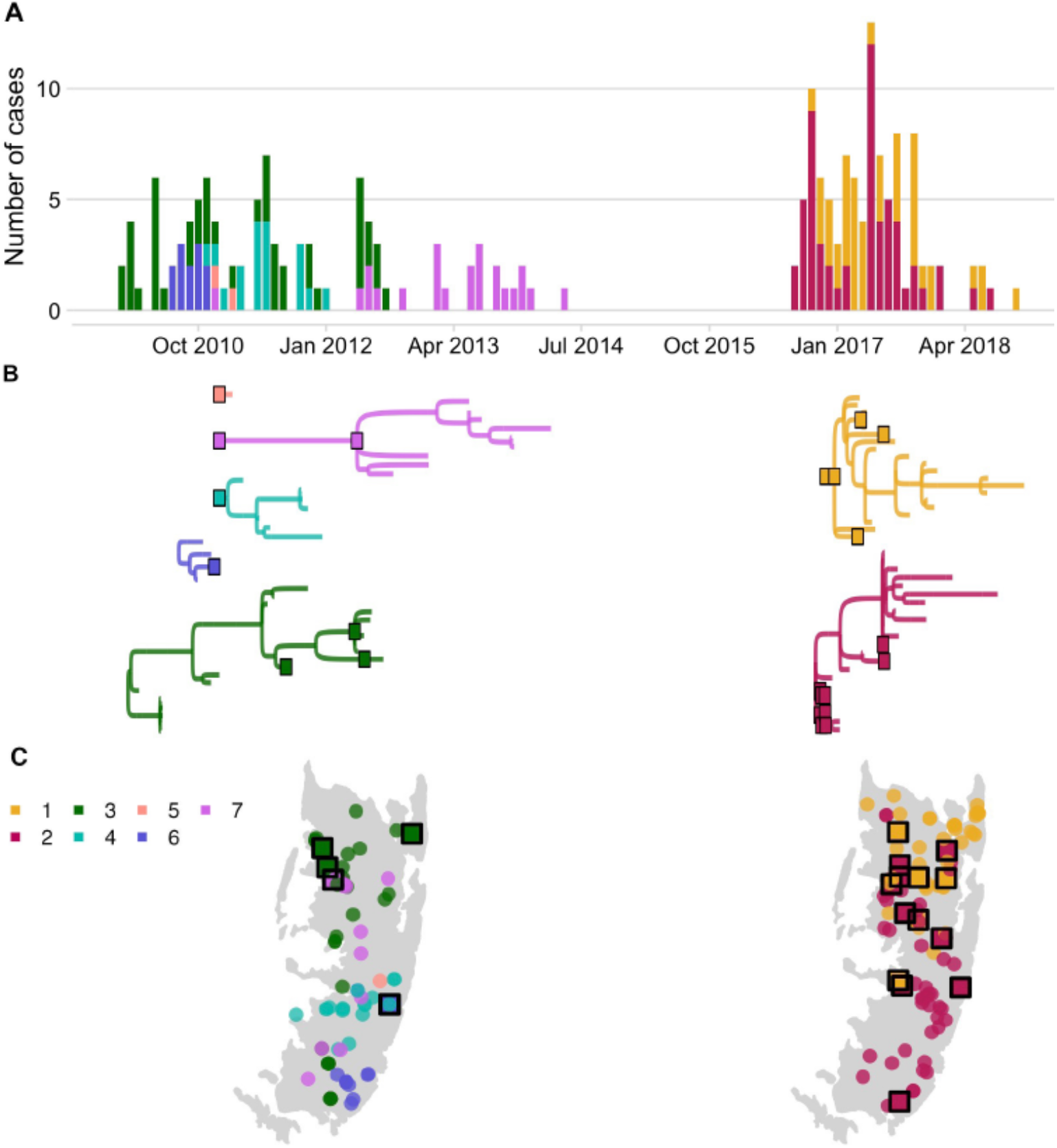
Rabies virus transmission chains inferred from epidemiological and phylogenetic data. **A)** Time series of cases coloured by their transmission chain. **B)** Consensus transmission tree (the highest probability transmission links that generate a tree consistent with the phylogeny) with chains pruned such that all unsampled cases are assigned to a sequenced chain of transmission. **C)** Spatial distribution of these cases over the two periods. In **B**, sequenced viruses from sampled cases are indicated by squares with a black outline, while only the tips are shown for unsampled cases. In **C**, unsampled cases are shown by a filled circle. In all panels, the data are colored by the transmission chain they were assigned to.

Of the bite patients presenting to the island’s four hospitals from 2010-2014 (n=117), a large proportion were bitten by probable rabid dogs (45-72% depending upon the status of unclassified biting dogs), while only a few patients that were bitten by apparently healthy dogs sought care during this period (6.6-12.8 per year, or 1.4-2.7/100,0000/year). Based on the probability of rabies progression following exposure^16^ and the occurrence of three human rabies deaths in 2010 (Table 1), we estimated that 10-31 rabid bite victims did not receive complete or timely PEP and through contact tracing we identified 21 such rabies exposures. The total rabies exposures that we detected from 2010-2014 (63-94) were within expectations from triangulating case detection and rabid dog behaviour^25^ (65 exposures, range 46-86), and consistent with a 0.66-0.89 probability of rabies-exposed bite victims receiving adequate PEP. During the 2016-2018 outbreak we traced 39 rabid bite victims who did not obtain adequate PEP (late and/or incomplete) and estimated that exposures received appropriate PEP with probability 0.72-0.78 (with the three deaths that occurred early in the outbreak suggesting around 10-31 rabid bite victims did not receive adequate PEP). Probable exposures per rabid dog were higher during the 2016-2018 outbreak than from 2010-2014 (1.3 vs 0.34-0.51, both adjusted for case detection) driven in part by variability in dog biting behaviour; two rabid dogs in 2017 each bit more than 10 people (Figure 2).

**Table 1.** Characteristics of probable human rabies deaths and reported reasons for inadequate Post-Exposure Prophylaxis (PEP).

| Year | Age | Bite site(s) | Type of wound | Reasons for not seeking PEP |
| --- | --- | --- | --- | --- |
| 2010 | 11-15 years | Both hands and the left palm | Severe wounds with broken bones | After first hospital visit, the child's family was not advised by health workers to return for subsequent PEP doses and family members were not aware of PEP requirements. |
| 2010 | >50 years | Lower left leg and upper thigh | Deep wounds with multiple tooth penetrations | Victim sought care at a facility (dispensary) that did not provide PEP. Received only first aid without referral to hospital for PEP. |
| 2010 | >50 years | Head (nose) and right arm | Lacerations to nose, large bite to arm, deep tooth penetration | After the wounds healed the victim did not seek their second or subsequent doses of PEP. |
| 2017 | 6-10 years | Neck | Large wound | PEP shortages in Pemba hospitals and prohibitive costs of seeking PEP elsewhere. |
| 2017 | 11-15 years | Face/head and shoulders | Severe wounds that led to hospitalisation | PEP shortages at the hospital where the victim was admitted. Health workers did not advise immediate PEP be sought from elsewhere. |
| 2017 | >50 years | Shoulders, legs and chest | Large wounds with deep tooth penetrations | Victim thought a single dose of PEP was sufficient for protection and ignored health worker advice to seek subsequent doses. |

Reasons reported for lack of, or inadequate, PEP varied (Table 1). No PEP shortages were reported whilst PEP was provided for free during the elimination demonstration project (2011-2014). But, at the start of the outbreak in late 2016 patients had to buy PEP (∼$12.9 per vial or >$38 for a complete course) and one child bitten in early 2017 by a confirmed rabid dog did not receive PEP due to a shortage. Although too late to be effective, health authorities sought PEP from Zanzibar when the child presented with symptoms but none was available. In desperation the family took the child to the mainland but with no rabies treatment options they were advised to return home where the child died on arrival.

Following the child’s death, Zanzibar’s Ministry of Health imported PEP and reinstated free-of-charge PEP provisioning. This policy change and sensitization around the outbreak likely contributed to increased health seeking, and understanding of the critical need for timely PEP. Contact tracing revealed that 17.5% of rabies exposures were not aware of the importance of PEP early on (2010-2014) compared to <4% during the outbreak (2016-2018) and similarly around 20% of rabies exposures early on (2010-2014), reported not being advised by health workers to obtain PEP, declining to 3% during the outbreak (2016-2018). Patients presenting for healthy dog bites also increased during the outbreak to ∼37/year (7.8/100,0000 vs 1.4-2.7/100,0000 previously).

Around $17,800 was spent on PEP for 542 bite patients who presented to hospitals in Pemba over the ten years. We estimated that this PEP prevented around 44 rabies deaths (95% confidence intervals: 27-59) among the 243 rabies-exposed patients who completed PEP, costing $405 per death averted. PEP cost-effectiveness could further increase to around $309 per death averted under the latest WHO recommendations for the abridged 1-week intradermal regimen.

Islandwide dog vaccination cost approximately $12,122 per campaign (Table 2, $13,145 for the campaign that reached most dogs) and interrupted transmission within four years of implementation, first in 2014 and again in 2018. Dog vaccination lapsed from 2014 allowing the two introductions in 2016 to spread widely. As three deaths occurred in both years when dog rabies was not under control (2010 prior to islandwide vaccination and 2017 before control of the outbreak), we speculate that over a ten-year time horizon without sustained dog vaccination, up to 30 rabies deaths might be expected on Pemba. Routine annual dog vaccination campaigns would prevent these expected deaths, at around $4,490 per death averted and would serve to keep Pemba rabies-free.

**Table 2.** Costs and cost-effectiveness of control and prevention activities. Exchange rate: 1 USD: 2296 Tsh (bank of Tanzania, 05/05/2022 https://www.bot.go.tz/). MoLDF = Ministry of Livestock Development and Fisheries, Tanzania; LTRA = Land transport regulatory authority; DoLD = Department of Livestock Development, Pemba. MSD = Medical Stores Department. LFO = Livestock Field Officer. *We do not include the cost of collecting dog vaccines from the airport. **each injection requires 5 minutes of health worker time and up to 8 injections for complete PEP.

| Intervention | Component cost description | Cost in USD |
| --- | --- | --- |
| PEP | Complete patient course | \$25.11 - 56.43 (ID vs IM) |
| | Average annual PEP provisioning | \$1485 |
| | Cost per death averted | \$405 (44 deaths averted from 2010-2020) |
| Mass dog vaccination | Cost per dog vaccinated | \$6.5 (range: 4.2-10.8, varied by campaign) |
| | Average annual islandwide dog vaccination campaign | \$12,122 (range: 11,493-13,145, depending on dogs vaccinated) |
| | Cost per death averted | \$4,490 (27 projected deaths averted over 10-years from initiating and sustaining islandwide vaccination) |
| One Health approach | Annual islandwide dog vaccination & PEP | \$13,607 - 12,998 (emergency PEP while transmission ongoing & precautionary PEP once eliminated) |
| | Cost per death averted | \$1,865 (71 deaths averted from both interventions) |

From 2019 onwards, in the aftermath of the outbreak when all transmission had been interrupted, approximately $876 was spent annually on PEP for patients presenting with bites from healthy dogs (Figure 1), i.e. precautionary expenditure post-elimination. Since dog vaccination interrupts transmission, we further estimate that eliminating rabies through dog vaccination would spare between 20 and 120 families each year on Pemba from rabid dog bites and the anxiety of needing to urgently obtain life-saving PEP. Precautionary PEP and sustained dog vaccination are necessary to mitigate ongoing risks from introductions. Thus over a ten-year time horizon, compared to the endemic situation, a One Health approach combining islandwide dog vaccination with emergency then precautionary PEP would avert approximately 71 deaths on Pemba, at a cost of $1,865 per death averted (Table 2), equitably tackling this preventable disease.

**<u>Movie S1. Rabies cases and inferred transmission chains on Pemba Island</u>**. Transmission reconstruction using the consensus links consistent with the phylogenetic assignments. Cases are animated each month, with animals that are incubating infections shown as empty circles until infectious when they transition to filled circles (note that many cases become infectious within the same month of exposure). Inferred transmission links are shown by curved lines and at the approximate time of the exposure event, coloured by transmission chain. Cases identified as introductions are designated by a filled square. The top panel shows the monthly time series of cases by transmission chain.

## DISCUSSION

Dog-mediated rabies is the quintessential zoonotic disease requiring coordinated public health and veterinary interventions as part of a One Health approach to end unnecessary suffering and deaths. Inequities in access to both human and animal vaccines manifest in the continued high burden of rabies in neglected communities around the world. Our study quantifies dog-mediated rabies transmission in an African setting, illustrating how rabies incidence in domestic dogs translates to human rabies exposures and how limitations in provisioning post-exposure vaccines results in human deaths. On Pemba, endemic rabies led to many exposures and three deaths in 2010, the year that our study began. Over the following four years consecutive islandwide dog vaccination campaigns were undertaken during a rabies elimination demonstration project. Initially, only low vaccination coverage was achieved (Figure 1), but prevention efforts improved after implementation challenges were overcome, including lack of dog vaccination experience and poor health seeking, conflated by expensive and inaccessible PEP. By 2014, transmission was interrupted. Unfortunately two independent introductions to Pemba in 2016, at a time when vaccination coverage was low, seeded a large outbreak causing three further rabies deaths in 2017. Attempts to respond locally were ineffective, until Pemba’s government re-established dog vaccination islandwide, after which rabies was rapidly eliminated. The island has remained rabies-free since October 2018.

Accumulating evidence illustrates how metapopulation dynamics maintain dog-mediated rabies via endemically co-circulating viral lineages.^6,7^ Despite Pemba being a relatively isolated island with a small dog population, genomic data revealed considerable RABV diversity, likely arising from historical introductions,^22^ and two contemporary introductions in 2016. Rapid outbreak spread from these introductions highlight the fragility of elimination. While dog rabies remains uncontrolled in nearby populations, reintroduction risks are high.^7,8,10,11,26^ Re-emergence is most likely if dog vaccination coverage is low, causing major public health and economic consequences.^8,11,27,28^ Introductions may be reduced through improved border control, but informal human-mediated movement of dogs is not easy to regulate. Scaling up coordinated dog vaccination should suppress the source of introductions and accelerate elimination, accruing and sustaining long-term benefits across much larger populations. While our study from a small island dog population represents a best case scenario, examples from Latin America show dramatic contractions of dog-mediated rabies when dog vaccination is scaled up and sustained. The last dog-mediated rabies foci on the continent remain only in very poor communities where dog vaccination has been inadequate.^3,9^

Our detailed contact tracing data from Pemba contrasts with very weak routine rabies surveillance in both humans and animals throughout much of Africa. Low case detection leads to underestimation of disease burden, lack of prioritisation, and difficulty ascertaining impacts of control, including whether disease has been eliminated, or is circulating undetected. The high case detection on Pemba generated confidence that elimination was achieved, twice.^29^ The subsequent re-emergence emphasises the need to maintain both surveillance and vaccination coverage where the risk of introductions from connected populations remains. The whole genome sequences generated in this study further revealed the underlying metapopulation dynamics of rabies circulation. By identifying introductions to Pemba and resolving their role in further spread it was possible to rule out sustained undetected transmission as the cause of re-emergence. Genomic approaches are increasingly affordable, and capacity for genomic surveillance is growing, accelerated over the course of the SARS-CoV-2 pandemic. For rabies, genomic approaches have potential to enhance the information that can be gleaned from routine surveillance and inform elimination programmes, which are likely to experience such introductions as they progress.

By tracing transmission within the dog population and from dogs to humans, we directly quantified lives saved by PEP showing how extremely cost-effective this intervention is, particularly when most bite patients present with rabies exposures rather than healthy dog bites. As an emergency medicine PEP is critical for rabies prevention. However PEP does not address the suffering caused from injuries inflicted by rabid animals, and on its own, is insufficient to sustainably protect the entire at-risk population. Our study shows how lack of awareness, expense and supply issues still prevent access to these emergency vaccines for marginalised populations. Only dog vaccination interrupts transmission in the reservoir so as to achieve the equitable goal of elimination. Although our crude cost-effectiveness analysis does not propagate uncertainties or fully disentangle the synergistic interactions of these interventions, the empirically-derived comparison illustrates the power of their combined delivery. Moreover, compared to other health interventions, a One Health approach for rabies is extremely cost-effective when translating into life years saved, even without considering recommended dose sparing intradermal PEP regimens or optimized delivery of dog vaccination^30^.

We conclude that the minimal investment needed for a One Health approach, to support access to life-saving emergency vaccines for humans and to achieve and maintain rabies freedom in source dog populations is exceptionally cost-effective and can bring rapid success. Lessons from Pemba should build confidence in the feasibility of eliminating rabies elsewhere on the African continent, given sustained commitment. Coordinated dog vaccination over sufficiently large scales will have the greatest and most long-lasting impacts in equitably tackling this preventable disease.

## Data Availability

Code to reproduce the analyses together with deidentified data are available from our public Github repository https://github.com/boydorr/Pemba_rabies.

## Acknowledgments

We are grateful to staff from the Animal and Health Departments on Pemba, Francois-Xavier Meslin from WHO HQ, Pelagia Muchuruza and Alphoncina Nanai from the WHO country office, and local community members for support. Aaron E. Lingo, Abdallah N. Mauly, Shamata S. Khamis, Mcha H. Mcha, Abdallah A. Mohamed, Hemed M. Ali and Abdallah M. Sudi all assisted with data collection and Daisy Jennings assisted with sequencing. The Tanzania Ministries of Health and Social Welfare, Livestock Development and Fisheries, the WHO Country Office-Tanzania, the National Institute for Medical Research, the Zanzibar Ministry of Health and Research Council, the Tanzania Veterinary Laboratory Agency, Afrique One-ASPIRE all provided permissions and collaborative expertise.

## Supplementary Methods

### Dog vaccination and coverage estimates

Pemba island (988 km^2^) situated fifty kilometres from the Tanzanian mainland is split into four administrative districts, with 121 villages (*shehias*) and a projected population of 438,765 in 2020 (Tanzania National Bureau of Statistics, 2012). Dog density is very low (1 dog to 118 humans), in this predominantly muslim population. Almost all dogs on Pemba are unconfined and 10-20% are estimated to be stray (unowned) potentially posing a problem for reaching coverage needed for elimination using central point strategies.

Four annual dog vaccination campaigns from 2011 to 2014 were organised as follows. One week before each campaign, a meeting was held between District Veterinary Officers, Livestock Field Officers (LFOs), and Community Animal Health Workers (CAHW) to review protocols and distribute vaccination equipment. CAHWs for each *shehia* then moved door-to-door inviting owners to bring their dogs to the nearest vaccination point and distributed posters. One day before the campaign, CAHWs walked repeatedly through each *shehia* announcing the forthcoming vaccination over a loudspeaker. Vaccination points were mostly situated in the centre of *shehias* but for small neighbouring shehias, vaccination points were located at central convenient locations. Each point was operated by two LFOs and a CAHW and campaigns ran from 9.00am to 3.00pm on a single day.

In 2016, a central point dog vaccination strategy was employed in *shehias* where dog rabies cases had been reported, as a response to control the outbreak. However, these efforts were not effective in preventing spread and remedial vaccination campaigns were undertaken, adopting a door-to-door strategy in some *shehias* where dog owners could not bring their dogs to allocated central points. Island-wide vaccination campaigns were conducted from 2017 onwards.

To estimate vaccination coverage at the *shehia* level through time, it was necessary to first estimate the concurrent dog population sizes. This was achieved using two datasets: 1) government dog population surveys for the years 2012 and 2017-2019, and 2) post-vaccination transects from the 2013-2014 vaccination campaigns, with associated total numbers of dogs vaccinated in the preceding campaigns. Where at least one collared (i.e. vaccinated) dog and >10 total dogs were observed during a transect, the dog population of a *shehia* at the time of the transect was estimated as:

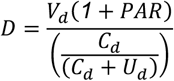

where *D* is the dog population size, *V*_*d*_ is the number of dogs vaccinated in the campaign preceding the transect, *C*_*d*_ is collared dogs, *U*_*d*_ is unmarked dogs, and *PAR* is the ratio of pups (<3 months) to adult dogs.^31^ *PAR* was estimated to be 0.256 from a census of the Serengeti District dog population in Northern Tanzania between 2008-2016.^32^ By multiplying by (1+*PAR*), we assume both that vaccination campaigns fail to reach pups, and that pups are not counted during transects.^31^

At least one Government or transect-based dog population estimate was available during the study period for each *shehia* on Pemba, with some *shehias* having estimates at up to six time points. For each *shehia*, the dog population in every month through the study period for which we did not already have an estimate was then projected. For months that lay between two known population estimates, a population projection was obtained via the exponential population growth rate calculated between those two estimates. For months where there was only a preceding or subsequent dog population estimate available, we projected the population based on a human:dog ratio calculated from this preceding/subsequent estimate and the human population size projected from the 2012 national census. In some cases, the projected dog population obtained for a month using this approach was lower than the number of dogs vaccinated during a vaccination campaign in that month. Where this occurred, the population estimates were adjusted as necessary to prevent coverage estimates exceeding 100%.

The coverage achieved by each vaccination campaign in each *shehia* was obtained by dividing the number of dogs vaccinated in that campaign by the estimated dog population for the month when the campaign occurred. We also estimated the monthly number of dogs with vaccine-induced immunity as follows:

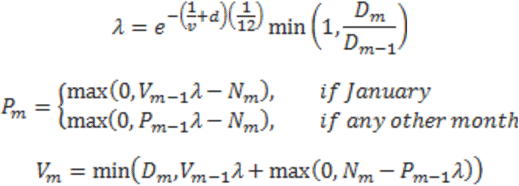

where *V*_*m*_ is the number of immune dogs at month *m, N*_*m*_ is the number of newly vaccinated dogs at *m, D*_*m*_ is the dog population at *m* estimated using the methods described above, and P_*m*_ is the number of immune dogs that were vaccinated during campaigns in previous years, not in the current year. Immunity wanes according to both *v*, the mean duration of vaccine-induced immunity (assumed to be 3 years), and *d*=0.595, the annual dog death rate.^33^ This approach conservatively assumes both that dogs that are immune from previous campaigns are preferentially vaccinated in subsequent campaigns and that, if the dog population declines between months, then this is a consequence of an above average death rate, rather than a below average birth rate. It also assumes that any top-up campaigns in a *shehia* in the current year focus on vaccinating susceptible dogs, avoiding re-vaccination of already vaccinated animals.

### Preliminary WGS phylogenetic analysis

Clade assignment in RABV-GLUE (main text) indicated that all Pemba sequences grouped within the RABV minor clade Cosmopolitan-AF1b. Therefore, an exploratory dataset of publically available genome sequences (coverage >90% of genome) from the Cosmopolitan-AF1b clade was obtained from RABV-GLUE (n=244) and supplemented with new sequences published in this paper (n=22, Table S1). An alignment was created in MAFFT^34^ and used to build a maximum likelihood (ML) phylogeny in IQ-TREE^35^ with default model selection. To simplify and focus analysis on Pemba outbreak cases, a subtree encompassing all 2016/17 Pemba sequences and relevant contextual sequences was extracted from the ML phylogeny and these sequences were used for Bayesian phylogeographic analysis in BEAST.^36^ In addition, one sequence from Uganda and one sequence from Rwanda were removed from the subset to avoid influencing phylogeographic analysis as the only two non-Tanzania sequences and two sequences (GenBank accessions: MN726823, MN726822) were removed as they contained a high proportion of masked bases (Ns) that affected tree convergence. This resulted in a reduced dataset of 153 sequences, exclusively from Tanzania, spanning the years 2001-2017. Note that this excluded two historical, previously published Pemba sequences (2010/12) belonging to a divergent lineage, previously defined as Tz5 in Brunker et al, 2015.^21^ TempEst was used to assess the temporal signal in the data, with a moderate association between genetic distances and sampling dates (R^2^=0.37) indicating suitability for phylogenetic molecular clock analysis in BEAST.^37^

### Partial genome phylogenetic analysis

RABV-GLUE was used to obtain an alignment of all sequences from the Cosmopolitan-AF1b minor clade up to the year 2017 (inclusive) regardless of genome position or length. This provided a much wider geographic and temporal context than only WGS sequences (Figure S1), enabling the placement of 2016/17 Pemba outbreak sequences into the context of known background diversity. The resulting dataset contained 2557 sequences from 21 countries (all sub-saharan Africa, aside from one Thailand sequence) from years 1980 to 2017 (Figure S1). This dataset was combined with the new sequences generated in this paper (n=22, Table S1) using MAFFT’s function to add new sequences to an existing alignment. R was used to categorise data into sequence types as follows: partial gene length sequences typically obtained from polymerase chain reaction (PCR) based diagnostic assays, full length (>90% coverage) gene sequences (gene) and whole (>90% coverage) genome sequences (wgs). Sequences were further categorised into genome position by gene: nucleoprotein (n), phosphoprotein (p), matrix protein (m), glycoprotein (g), RNA polymerase (l). This facilitated detailed exploration of publicly available RABV sequence data to obtain the most informative datasets to compare Pemba outbreak sequences.

**Figure S1.**
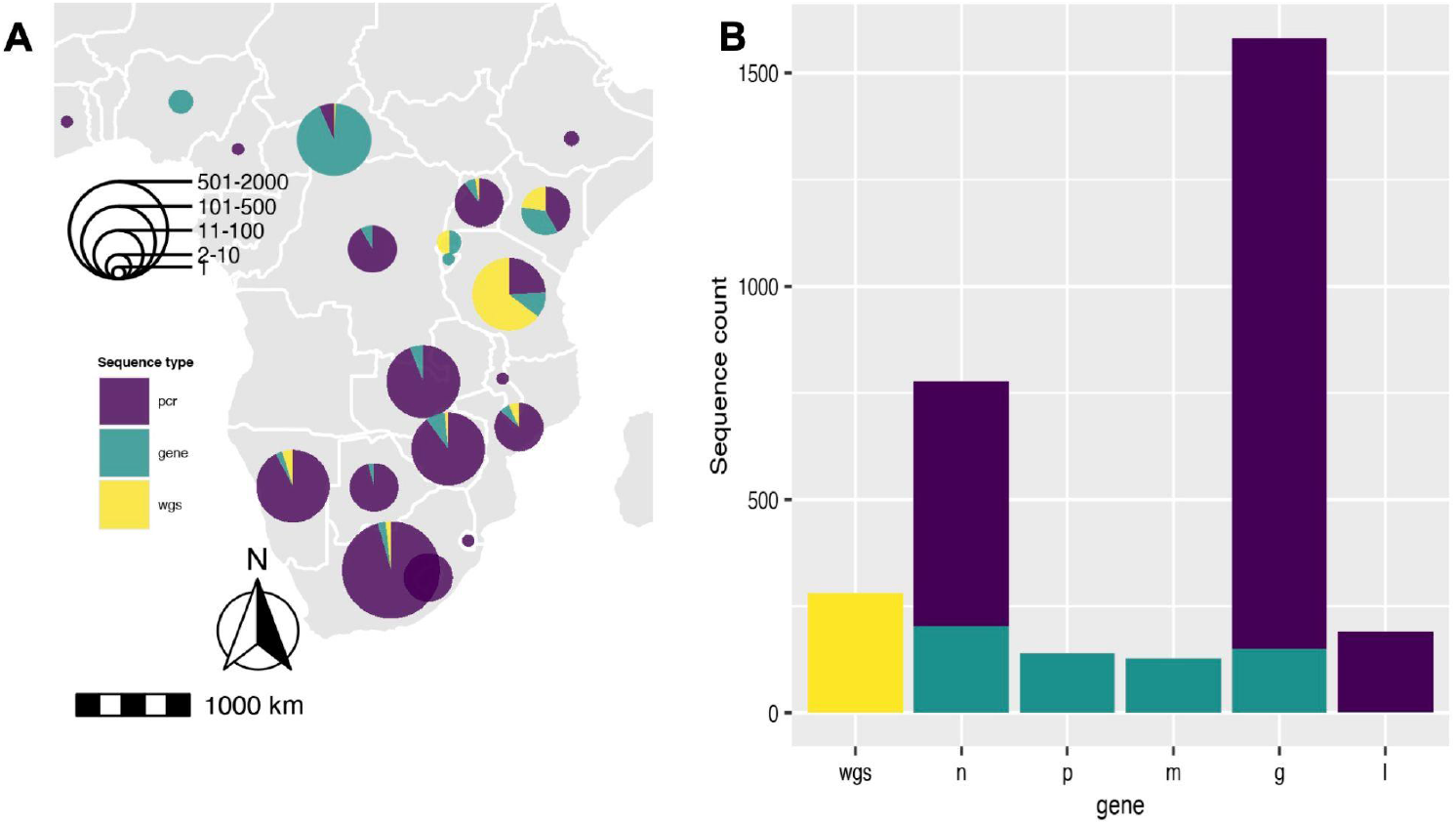
Rabies virus sequences within the Cosmopolitan-AF1b minor clade. Sequences obtained from RABV-GLUE (n=2557) **A)** Spatial distribution of sequences across Africa categorised by type: partial gene length sequences typically from polymerase chain reaction diagnostics, full length (>90% coverage) gene sequences (gene) and whole (>90% coverage) genome sequences (wgs). **B)** Number of sequences per gene (pcr or whole gene) or whole genome. Nucleoprotein (n), phosphoprotein (p), matrix protein (m), glycoprotein (g), RNA polymerase (l).

N and G genes were the most commonly sequenced, with 1042 and 1876 sequences respectively (including WGS), and both gene datasets were used to contextualise Pemba sequences. There was wide variation in the portion of each gene covered and the length of sequences (N: from 203 to full length 1353 basepairs (bp); G: from 277 to full length 1575bp). The N gene dataset constituted a wider and more even geographic distribution (18 countries) whereas the G gene (16 countries) data was predominantly from South Africa and Tanzania. Background ML phylogenies were produced in IQ-TREE with default settings, using alignments of the variable length gene sequences. Extreme outliers with long branches (upper and lower 1st percentile of branch length distribution) were removed and a subtree extracted stemming from the most recent common ancestor (or one node back from) of all Pemba (historical and outbreak) sequences.

Sequences from these subtrees (N and G gene) were subject to a more robust phylogenetic reconstruction with rapid bootstrapping in IQ-TREE and an outgroup sequence from the Cosmopolitan-AF1a minor clade (GenBank Accession: KC196743). Bootstrap values were generally low across the phylogenies, most likely due to the use of short, variable length sequences and therefore should be interpreted with caution. We ‘zoomed in’ on clusters within these subtrees to identify the closest relatives to Pemba outbreak cases, Figure S2. For Pemba cluster 1 (Figure 2b, Figure S2a &b), sequences were most closely related to sequences from the Serengeti District in northern Tanzania according to both N and G gene datasets. Whereas Pemba cluster 2 sequences (Figure 2c,Figure S2c &d) share a common ancestor with N gene sequences from Zanzibar, a neighbouring island, from the same period (2016/17). This suggests a possible link between rabies outbreaks on these islands and/or a common source of introduction.

**Figure S2.**
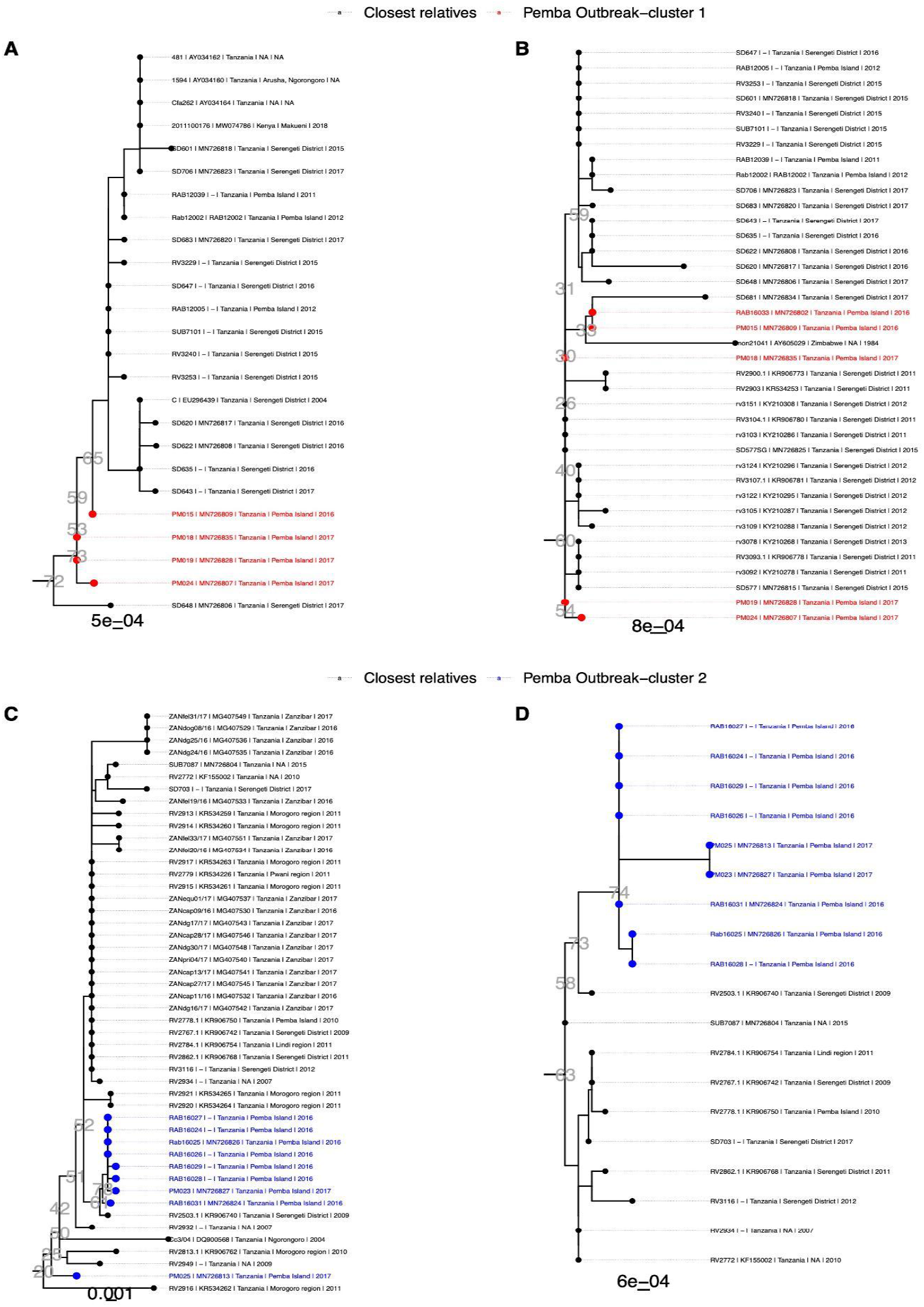
Phylogenetic clusters of RABV sequences from nucleoprotein (N) and glycoprotein (G) datasets showing the closest relatives to Pemba outbreak cases. Subtrees extracted from background phylogenies constructed from datasets of N (n=1042) or G sequences (n=1876). **A)** & **B)** Pemba cases from cluster 1 (red), shown in main text Figure 2b, in relation to N and G gene sequences respectively; **C)** & **D)** Pemba cases from cluster 2 (blue), shown in main text Figure 2c, in relation to N and G gene sequences. Tip labels indicate metadata in the format <isolate id | GenBank accession | country | location | year>. Bootstrap values are shown next to nodes of interest in grey text and scale bars represent the number of substitutions per site.

### Transmission chain reconstruction

Using the case data, we reconstructed transmission trees probabilistically as previously described.^6^ We refined the tree-building algorithm to generate trees consistent with the phylogeny. First we generated a pairwise SNP distance matrix from the alignment using the ape package^38^ in R from which to assign independent lineages. We then built a directed graph of the transmission tree (Figure S3A) identifying edges connecting mismatched phylogenetic lineages. We sampled an edge for each of these paths to break, first by frequency (i.e. how often they occur in paths with mismatches), then by the scaled probability of the spatiotemporal distance to the assigned progenitor, generally selecting lower probability links to resample (Figure S3B). For edges that were broken, we sequentially resampled a progenitor from those that generated trees consistent with the phylogenetic assignments (Figure S3C and D).

**Figure S3.**
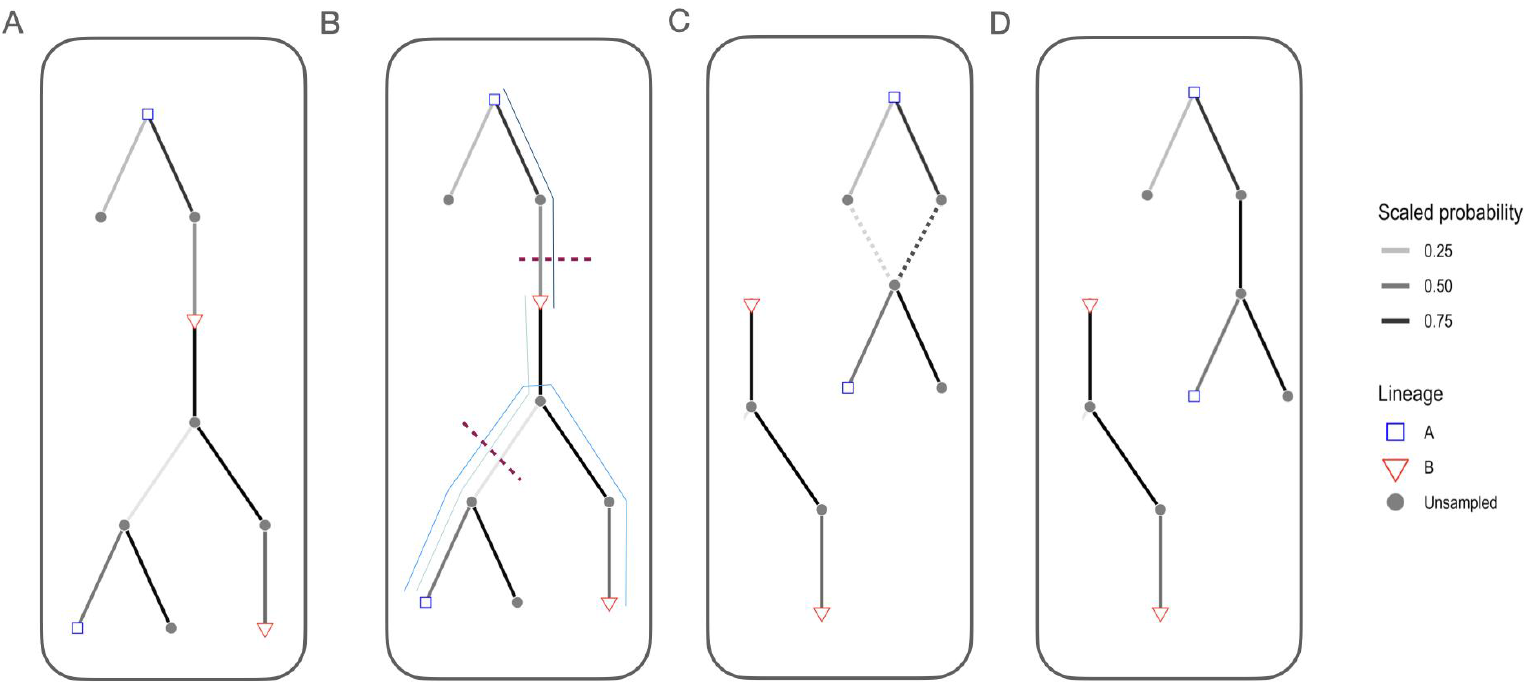
Steps for building transmission trees consistent with phylogenies. **A)** We first assigned progenitors probabilistically based on the serial interval distribution and dispersal kernel, as previously described^6^ and built a directed graph based on this tree. **B)** We then identify the paths between mismatched edges that are inconsistent with phylogenetic assignments (thin blue lines) and select the links to break first by filtering to the most frequently occuring edges before selecting one edge per path by the scaled probability of the link (selected links to remove are indicated by the red dashed lines, note that the same edge can be removed to resolve multiple phylogenetic mismatches). **C)** For cases where links were removed, we reassign progenitors sequentially (in a random order), rescaling the probability to only those progenitors that can generate phylogenetically consistent trees (dashed lines). If no phylogenetically consistent progenitor is identified the case becomes the index case of the transmission chain. Panel **D)** shows the final tree with phylogenetically consistent links between cases.

To further resolve transmission chains we applied additional pruning steps to filter out case pairs where the time interval or distance exceeded the 99th percentile of the reference distributions for the serial interval and distance kernel distributions. The tree reconstruction methods are wrapped into an R package, available at github.com/mrajeev08/treerabid and archived on Zenodo (DOI: 10.5281/zenodo.5269062). We compared resulting pruned trees (split into transmission chains) to the single unpruned tree, which connected all cases together, and considered transmission trees reconstructed only from the epidemiological data versus those pruned to be consistent with the phylogeny. The uncertainty in progenitor and lineage assignments according to pruning approaches and with inclusion of genetic data are shown in Figure S4. For each pruning algorithm we compared across the consensus trees (i.e. the most frequently assigned progenitors for each case), the Maximum Clade Credibility (MCC) trees (the tree within the bootstrap that had the highest product of progenitor probabilities) and the majority transmission trees (the tree within the bootstrap that had the highest number of consensus progenitors), shown in Figures S5, S6 and S7 respectively. We examined the relationship between mean secondary cases (Re) in relation to vaccination coverage at the time of symptoms in the shehia where each case occurred (Figure S8).

**Figure S4.**
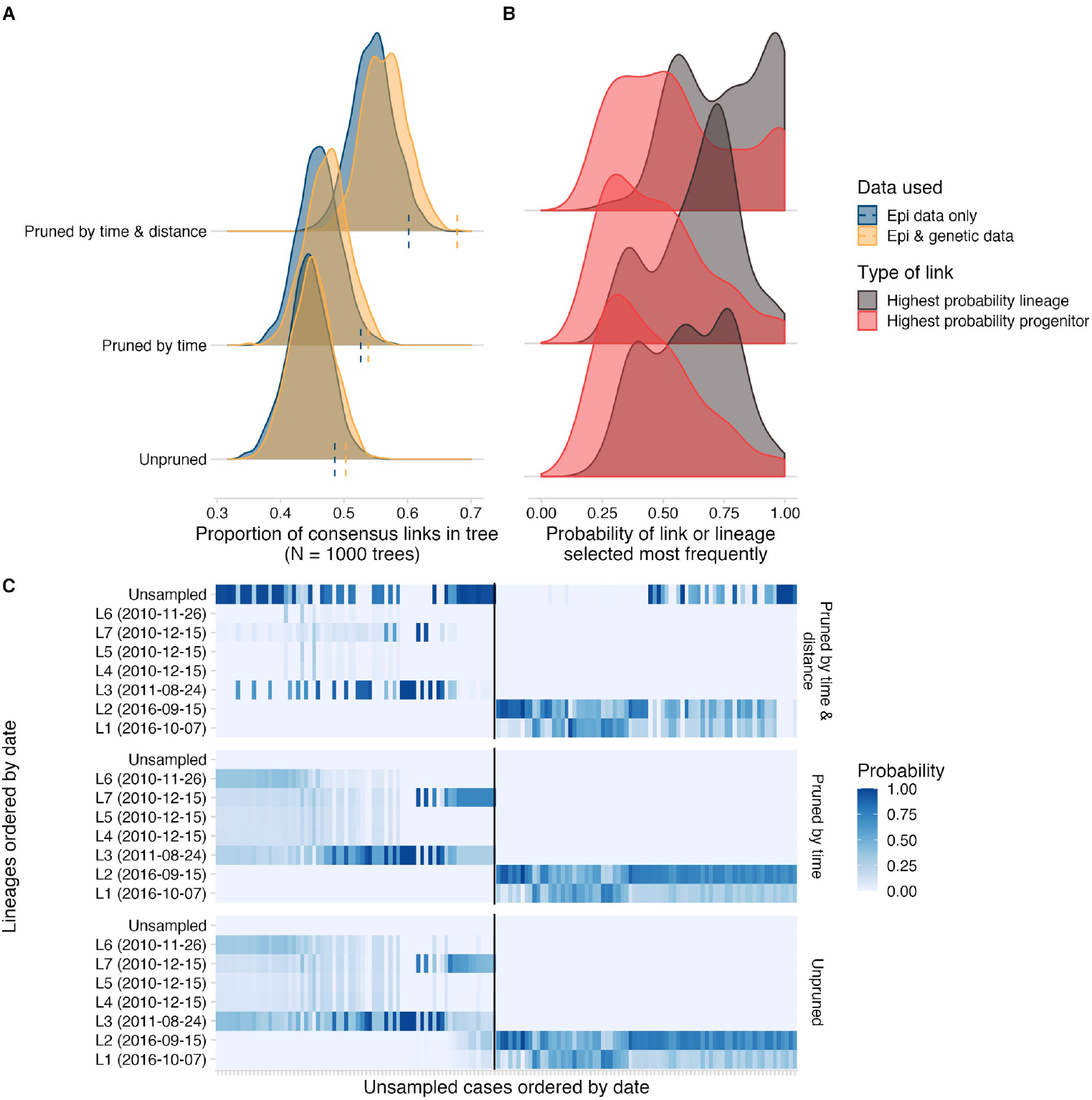
Comparing pruning approaches for transmission tree reconstruction. **A)** The distribution of topological uncertainty (that is the proportion of consensus links, or the most frequently selected progenitors in each tree in the bootstrap) for approaches pruning by time and/or distance thresholds (y-axis) and using epidemiological data only vs. integrating phylogenetic data. **B)** For approaches integrating genetic data, the comparison of the consensus progenitor probabilities (i.e. the proportion of times the most frequent progenitor is selected for each) vs. phylogenetic probabilities (the proportion of times a case is assigned to the most frequent lineage). **C)** For approaches integrating genetic data, the probability at which each unsampled case (x-axis, ordered by the date of symptoms) is assigned to a sampled lineage (y-axis ordered by date of symptoms; chains that were entirely unsampled are at the top). The black line separates pre-2015 cases from those that occurred after i.e. from the outbreak that began in 2016.

**Figure S5.**
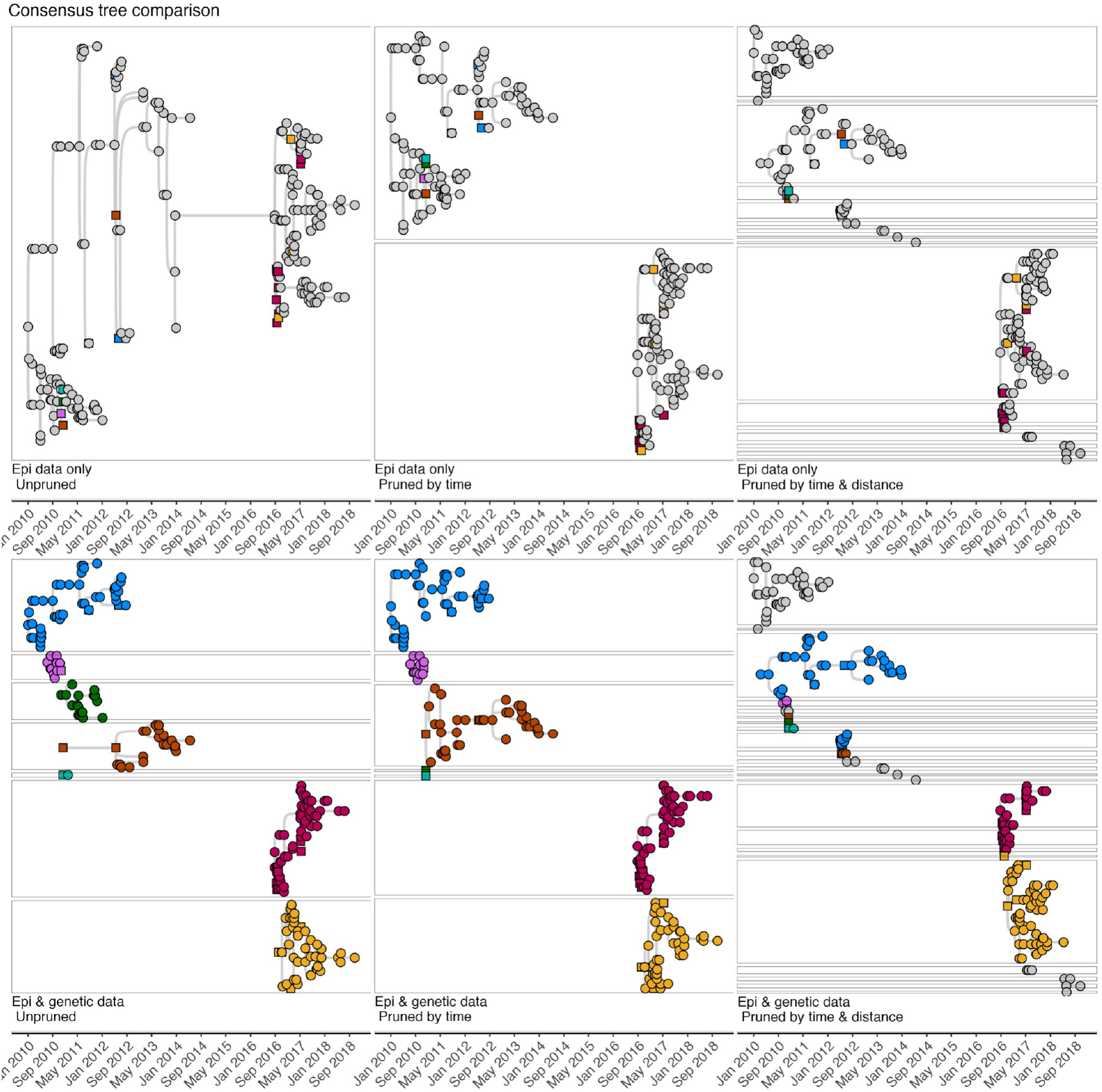
Comparison of the consensus transmission trees. (i.e. the most frequently assigned progenitors for each case) across pruning algorithms. Squares indicate cases that were sampled and their viruses sequenced and circles indicate cases without sequence data.

**Figure S6.**
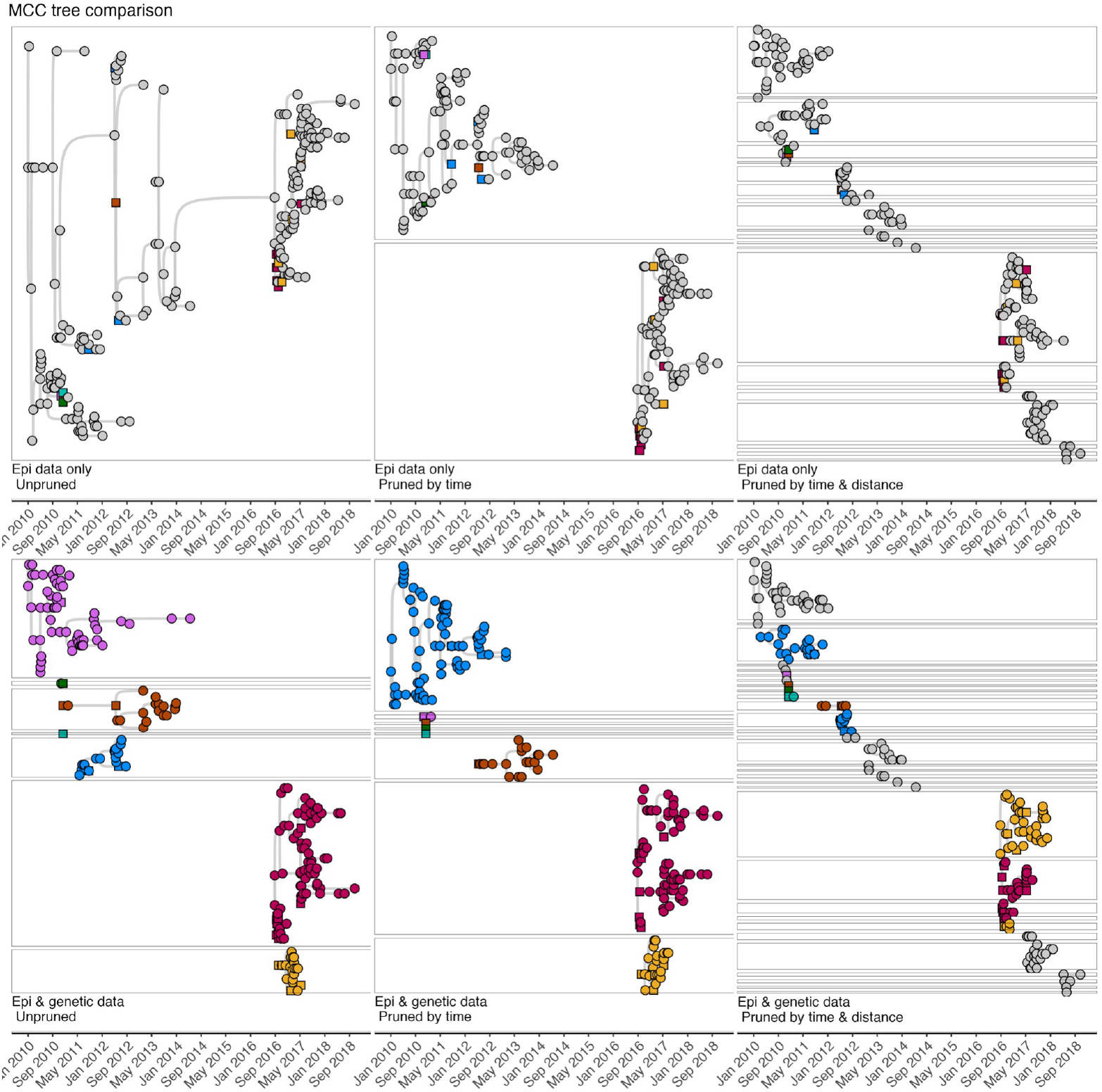
Comparison of the maximum-clade credibility (MCC) trees. (the tree within the bootstrap that had the highest product of progenitor probabilities) across pruning algorithms. Squares indicate cases that were sampled and their viruses sequenced and circles indicate cases without sequence data.

**Figure S7.**
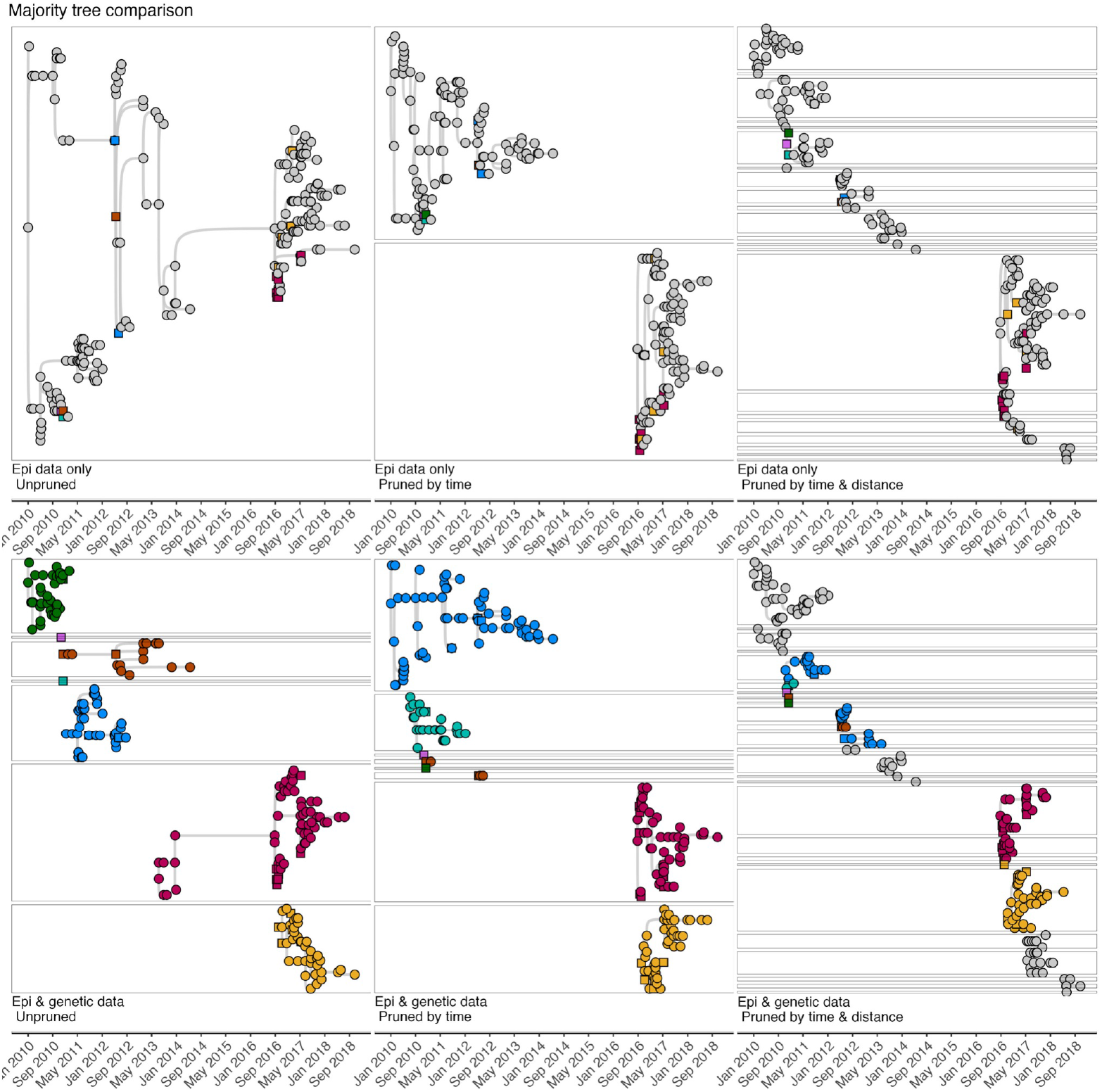
Comparison of the majority trees. (the tree within the bootstrap that had the highest number of consensus progenitors) across pruning algorithms. Squares indicate cases that were sampled and their viruses sequenced and circles indicate cases without sequence data.

**Figure S8.**
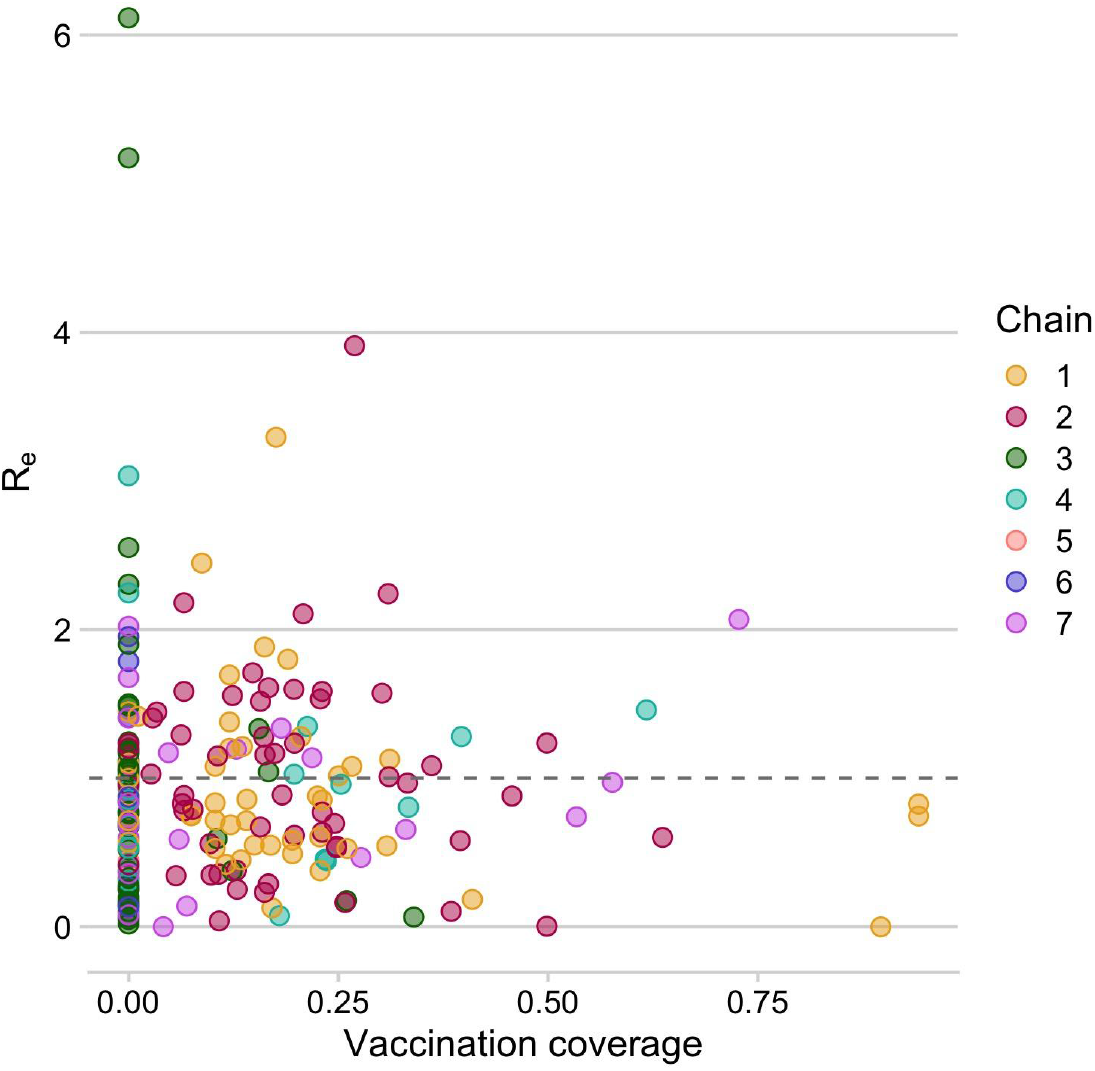
Vaccination coverage versus inferred R for each case. Number of secondary cases resulting from each case was averaged across the bootstrapped set of transmission trees generated by pruning to be consistent to the phylogeny. The points are the mean values for each case colored by their consensus assignment. Vaccination coverage is estimated at the time of symptoms in the *shehia* where the case occurred. The grey dashed line indicates an R of 1.

Using recently developed analytical methods^6,39^ we estimated the level of case detection from our contact tracing. As previously described, we used the observed time between statistically-linked cases (from the transmission tree reconstruction) and the serial interval distribution for rabies, to fit the simulated distribution of numbers of unobserved intermediates, implicitly assuming all infected individuals have the same probability of being detected. To account for the long-tailed distribution of serial intervals, we sorted simulated values for initial intervals to most closely match observed values (i.e. so long incubators are accounted for and not always taken to be cases with multiple generations separating them from their progenitors). This approach with sorting generally performs better than the unsorted approach,^6^ but tends to underestimate detection probabilities by about 10%, in particular for values between 0.3 - 0.75 (Figure S9A). We then examined the fit across a range of detection probabilities for the endemic period (2010-2014), the subsequent outbreak (2016-2018) and overall (Figure S9B), applying the method to 100 bootstrapped trees generated by the pruning strategies described above, and to the majority tree and the MCC tree, taking the mean of 10 estimates as the detection probability for each tree. We also confirmed that the method was robust even for small chains of transmission, given detection of at least one case (Figure S9C). We compared estimates according to alternative tree pruning algorithms, and with and without genetic information, finding that estimates were robust (Figure S10).

**Figure S9.**
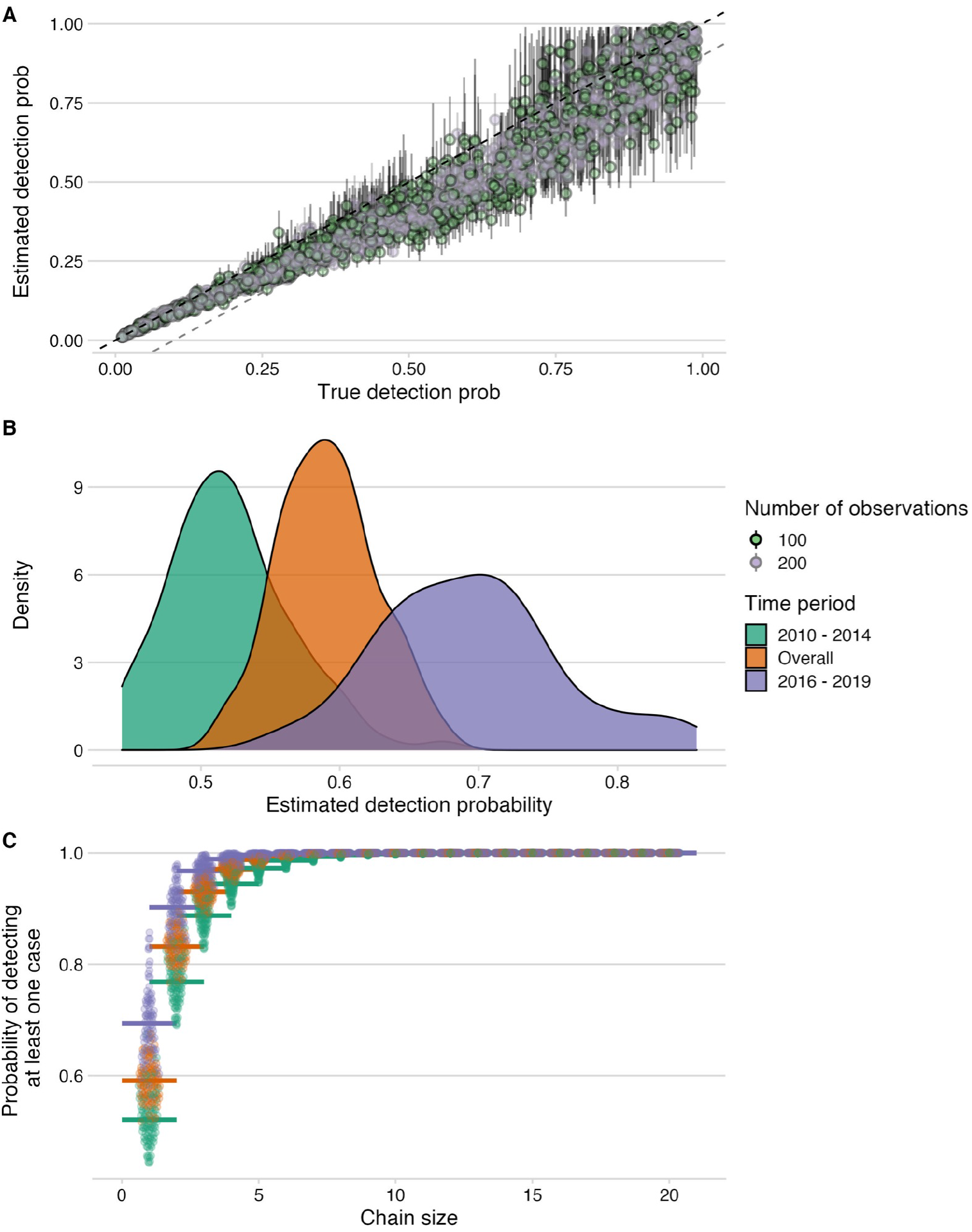
Estimation of detection probabilities. **A)** Estimated detection probabilities from simulated times between linked cases given a known detection probability (x-axis). Colours indicate the number of detected cases used in the simulations. The points show the mean and the lines the range of 10 estimates per simulation. The black dashed line shows the 1:1 line and the grey dashed line the 1.1:1 line. Estimates of detection from these simulations are generally recoverable, although with smaller sample sizes, the estimates are more dispersed. **B)** Detection probabilities estimated from times between linked cases using the tree algorithm with pruning by the phylogenetic data only. For the estimation, the times between linked cases for a subsample of bootstrapped trees (N = 100), as well as the MCC and the majority tree were used. The colours indicate the time period for which estimates were generated, 2010-2014 (pre-elimination), 2016-2018 (reemergence) and overall combining cases. **C)** Probability of detecting at least one case given estimated detection probabilities and chain sizes (x-axis) with colours corresponding to the period for which estimates were generated.

**Figure S10.**
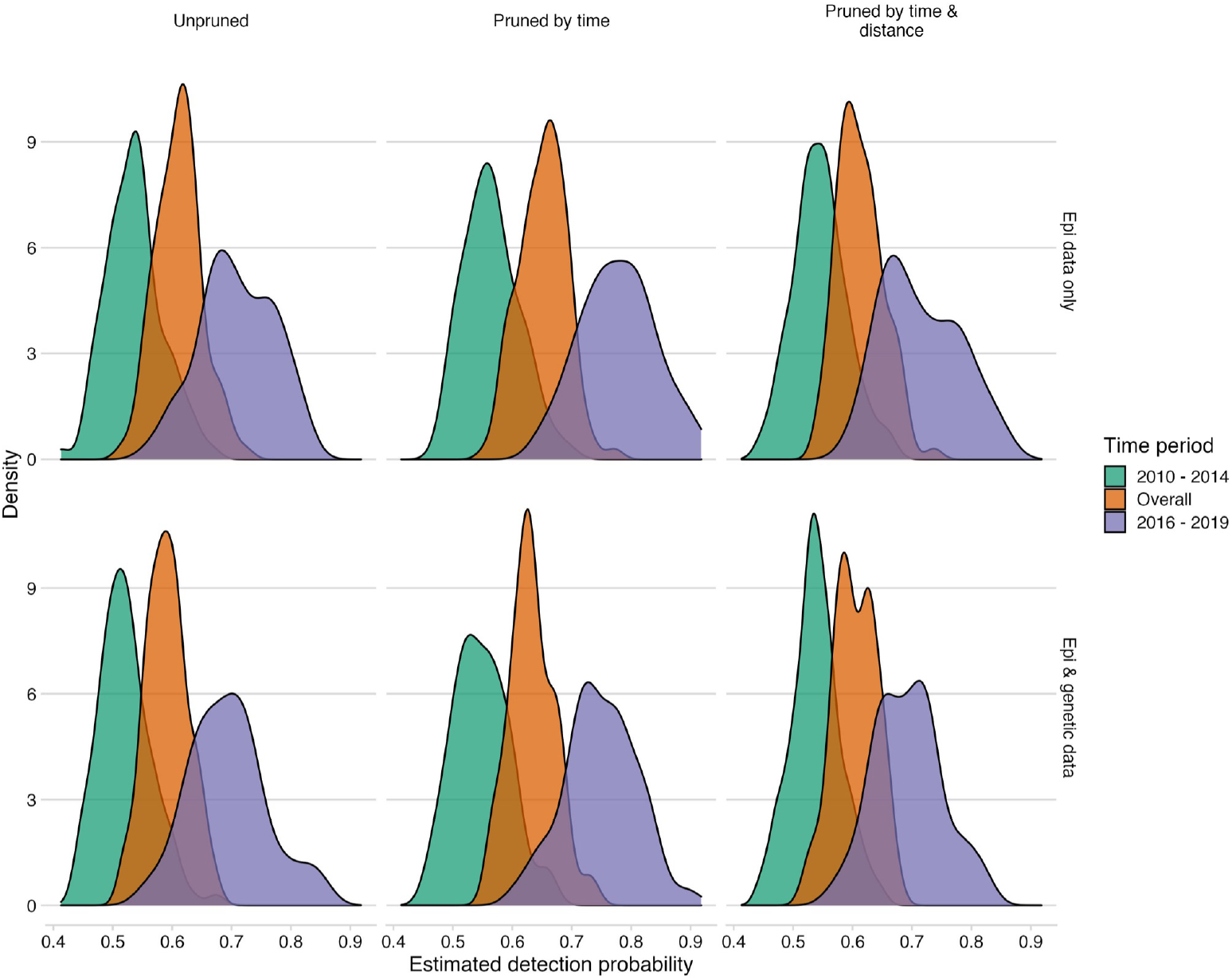
Comparison of detection estimates across pruning algorithms. For the estimation, the times between linked cases for a subsample of bootstrapped trees (N = 100), as well as the MCC and the majority tree were used. The colours indicate the time period for which estimates were generated, 2010-2014 (the pre-elimination period), 2016 - 2019 (the reemergence period) and overall combining cases.

## Supplementary Tables

**Table S1.**
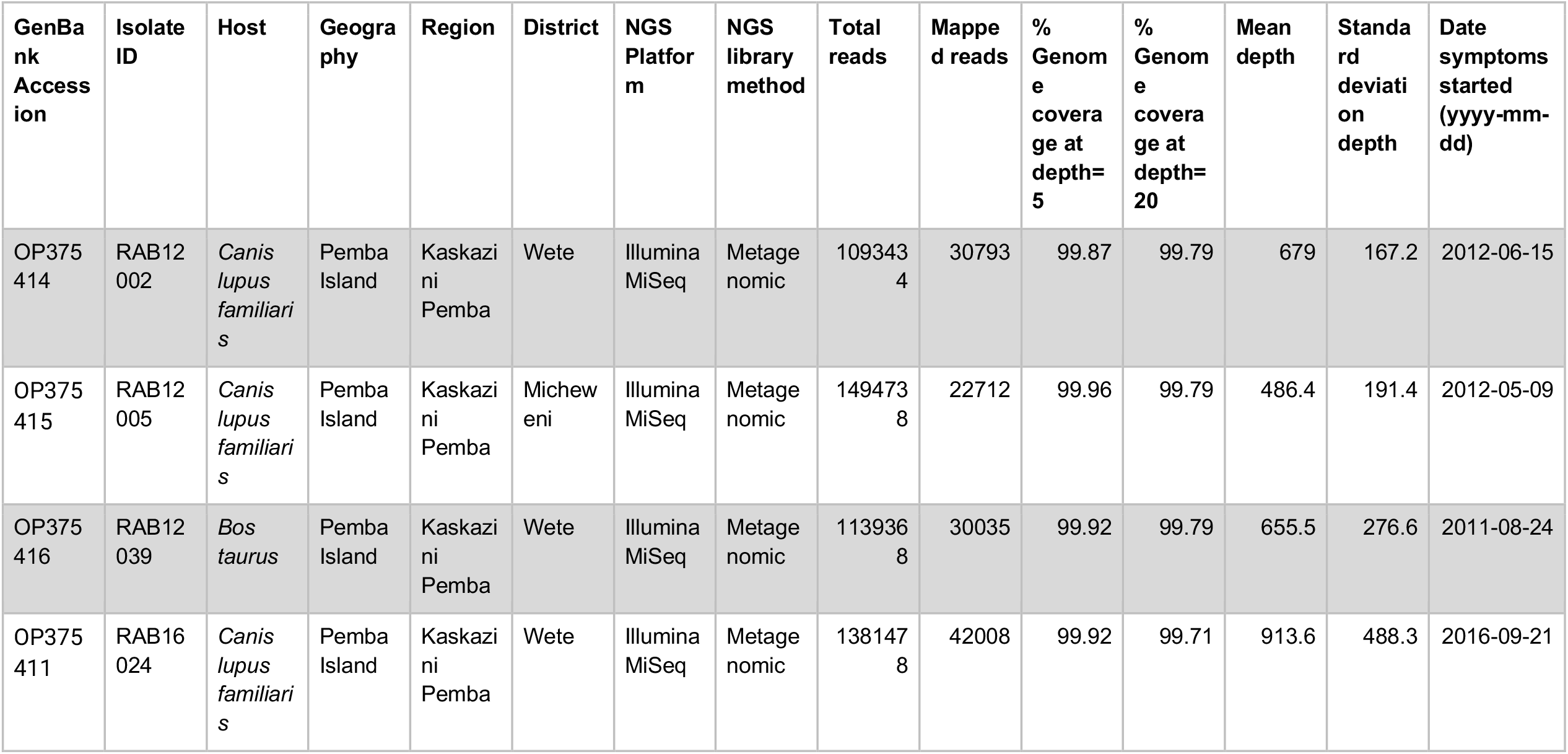
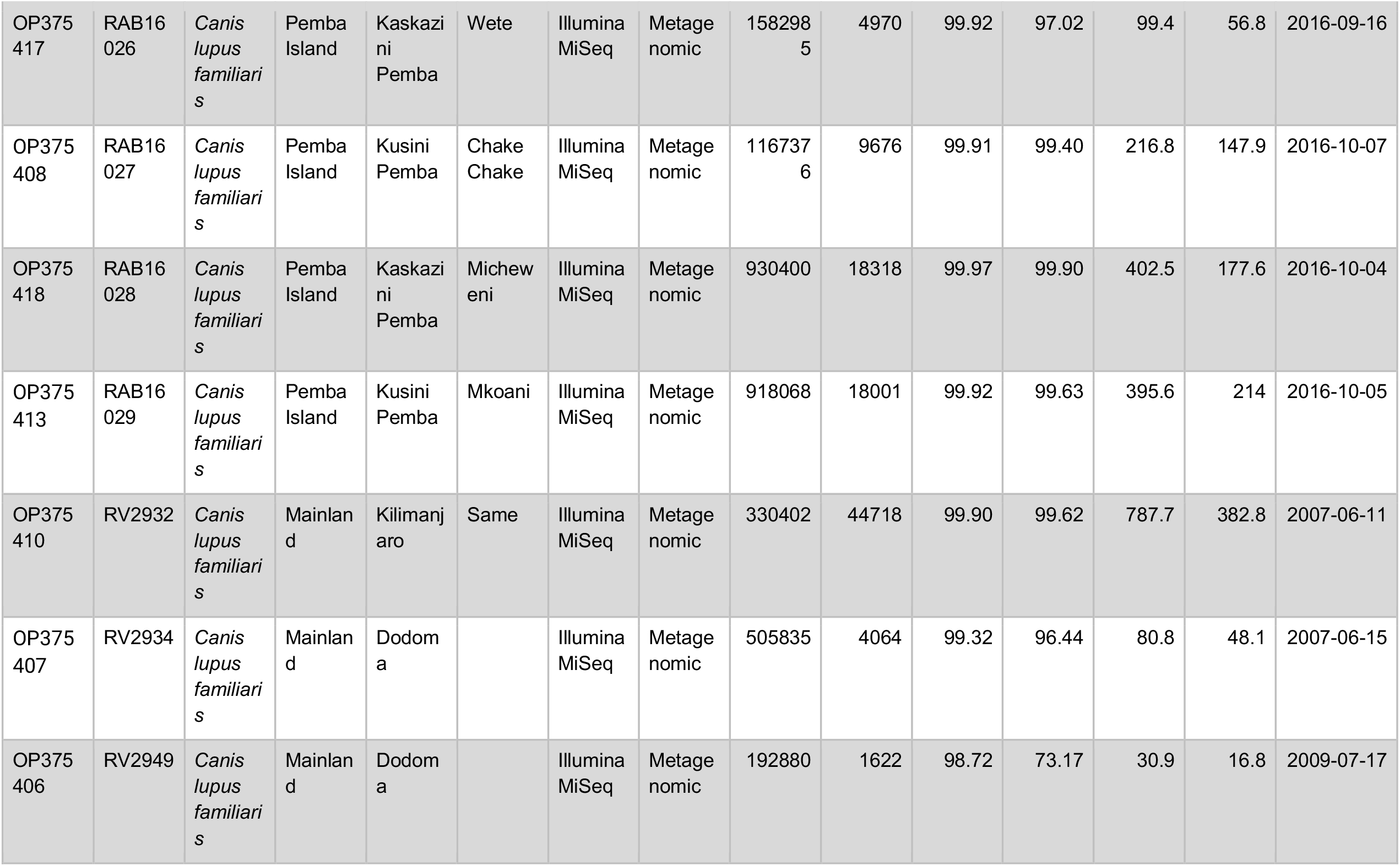
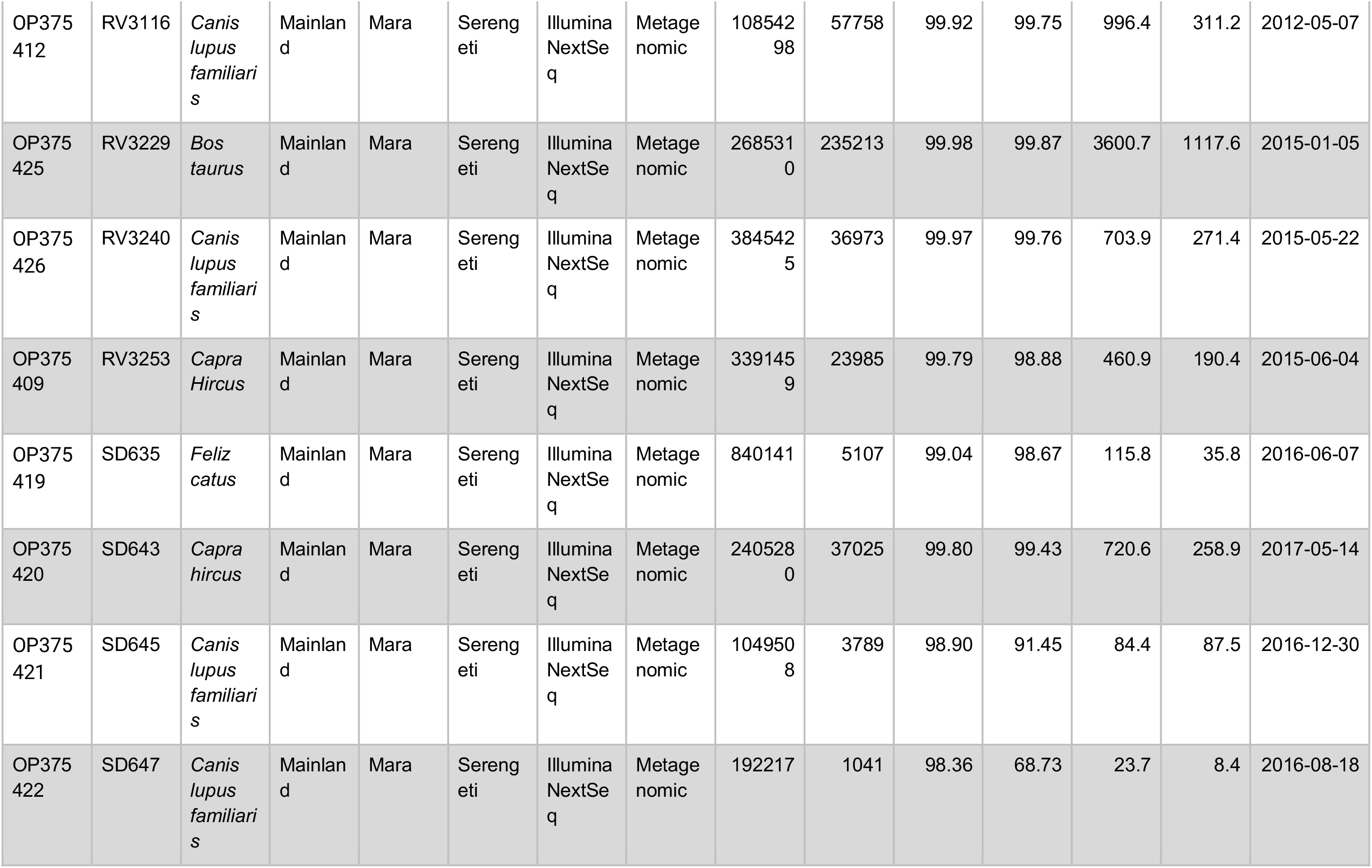
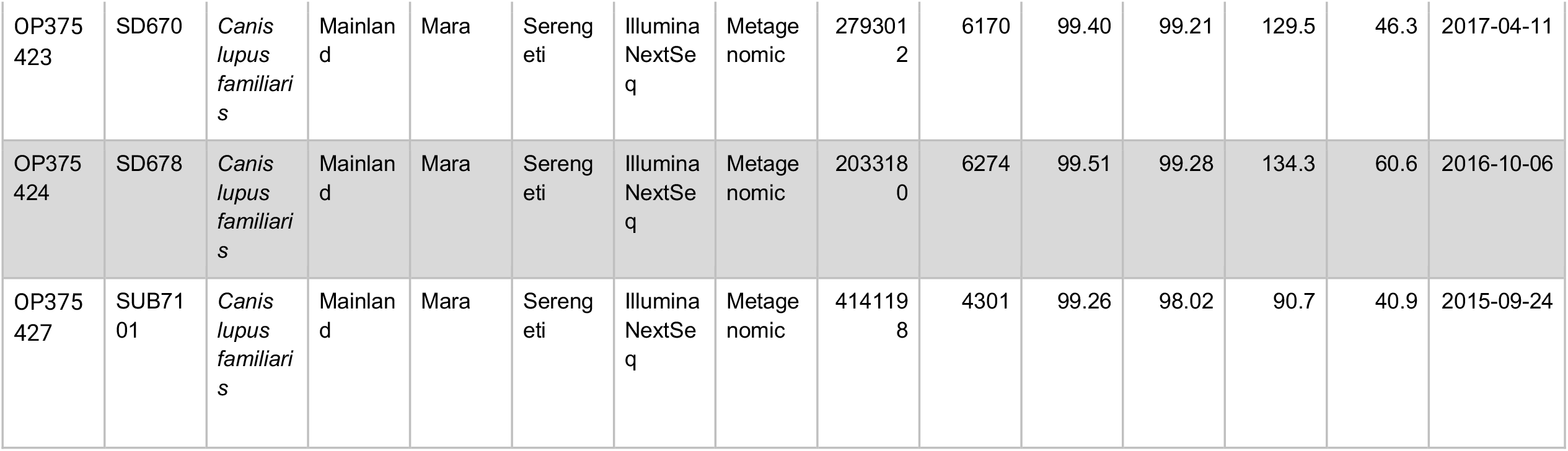
Epidemiological and next generation sequencing (NGS) metadata for 22 rabies virus isolates newly sequenced for this study.

**Table S2.** Costs of rabies control and prevention activities. Exchange rate: 1 USD: 2296 Tsh (bank of Tanzania, 05/05/2022 https://www.bot.go.tz/). MoLDF = Ministry of Livestock Development and Fisheries, Tanzania; LTRA = Land transport regulatory authority; DoLD = Department of Livestock Development, Pemba. MSD = Medical Stores Department. LFO = Livestock Field Officer. *We do not include costs of vaccine collection from the airport. **each injection requires 5 minutes of health worker time and up to 8 injections per PEP course.

| Intervention | Cost variables | Unit Cost (USD) | Number | Source |
| --- | --- | --- | --- | --- |
| Mass dog vaccination | Dog vaccine | 0.65 | Per dog | MoLDF |
|  | Consumables (syringes, needles) | 0.05 | Per dog | MSD price catalogue 2022/23 |
|  | Stationary (registers, certificates) | 4.36 | Per district | Local prices |
|  | Advertising for campaigns | 7.45 | Per vaccination day | DoLD |
|  | Transport (fuel) for team* | 7.45 | Per central point | DoLD |
|  | Assistant allowance | 2.18 | Per vaccination day | DoLD |
|  | LFO allowance | 13.07 | Per vaccination day | DoLD |
| Post-exposure vaccination | Consultation & wound care | 10.9 | Per patient | National health Insurance scheme |
|  | Post-exposure vaccine | 10.98 | Per vaccine vial | MSD price catalogue 2022/23 |
|  | Health worker time | 2.11** | Per patient | Tanzania Public service management and good governance |

